# Association between Preserved Ratio Impaired Spirometry with mortality and long-term cardiovascular outcomes in Chinese adults

**DOI:** 10.1101/2025.02.04.25321451

**Authors:** Mengya Li, Yang Li, Mengxin Chen, Duong Mylinh, Qiujing Cai, Biyan Wang, Sumathy Rangarajan, Kai You, Jiying Li, Salim Yusuf, Wei Li, Zhiguang Liu, Bo Hu, Lap Ah Tse, PURE-China Investigators

## Abstract

**Background:** To examine the prevalence and characteristics of PRISm among Chinese individuals, as well as its association with future mortality and cardiovascular (CVD) outcomes.

**Methods:** This was a substudy of the Prospective Urban Rural Epidemiology (PURE) study, which recruited 40,279 individuals aged 35-70 years from 115 urban and rural communities in 12 provinces across China between 2005 and 2009. At baseline, participants were categorized into subgroups based on PRISm, airflow obstruction (AO), and normal spirometry. Follow-up was conducted every three years to obtain information on major cardiovascular events and mortality. Cox frailty proportional hazard regression was used to estimate the hazard ratios (HR) and 95% confidence interval (95%CI).

**Results:** The baseline prevalence rates of PRISm, AO and normal spirometry were 29.3%, 8.5%, and 62.2% respectively. Over a median follow-up period of 11.9 years, 2,214 deaths, with 773 attributed to CVD, and 3,507 major CVD events were observed. After adjusting for potential confounders, individuals with PRISm, comparing to those with normal spirometry, exhibited significantly elevated risks of all-cause mortality (HR 1.42, 95%CI [1.29, 1.58]), CVD mortality (HR 1.35, 95%CI [1.14, 1.60]), major CVD events (HR 1.16, 95%CI [1.07, 1.25]), myocardial infarction (HR 1.34, 95%CI [1.15,1.56]), and heart failure (HR 2.02, 95%CI [1.46, 2.79]).

**Conclusions:** PRISm, a frequently observed result in spirometry among the general Chinese population, exhibits a substantial association with long-term all-cause mortality, CVD mortality, major CVD events. Further investigation is warranted to comprehensively compared the underlying pathophysiologic connection between PRISm and CVD as well as mortality.

**What is already known on this topic:** PRISm is considered a transient state with higher transition rates to both normal and obstructive spirometry, often indicating progression to COPD, which is associated with increased respiratory symptoms, diminished quality of life, and a higher risk of cardiovascular events and all-cause mortality. Given the limited understanding of PRISm, there are several studies on the association between PRISm and health outcomes have been carried out in different populations since the concept of PRISm was introduced in 2014. Longitudinal studies based on population cohorts such as COPDGene, UK Biobank, Rotterdam, and NHLBI have shown that PRISm is associated with an increased risk of all-cause mortality and adverse cardiovascular outcomes, with similar results found in a study based on a Japanese population. Lung function differs substantially between races and regions. Most of the existing PRISm-related studies are based on populations in developed countries such as Europe and the United States, and the conclusions of these studies should not be directly generalized to East Asian populations, including China. Currently, there is only two studies based on a Japanese and Korean aimed at exploring the relationship between PRISm and all-cause mortality and adverse cardiovascular outcomes, but the sample size of these study is relatively small, and the statistical power of the conclusions is relatively limited.

**What this study adds:** This is the first longitudinal study examining the association of PRISm with the risk for all-cause mortality and adverse cardiovascular outcomes in a general Chinese population. The results showed that the prevalence of PRISm in the general Chinese population is 29.3%, which is significantly higher than previous studies. Compared to individuals with normal spirometry, the population with baseline PRISm had a significantly increased risk of all-cause mortality, CVD mortality, myocardial infarction (MI), and heart failure (HF), and showed a trend towards higher risk than those with baseline airflow obstruction (AO, although no statistically significant difference was observed).

**How this study might affect research, practice or policy:** Our findings support that the early prevention, diagnosis, and intervention of PRISm may offer an important strategy to alleviating the high CVD burden in China.

## Introduction

Preserved Ratio Impaired Spirometry (PRISm) is now a recognized spirometric pattern, characterized by a reduced forced expiratory volume in one second (FEV_1_) and a normal FEV_1_ to forced vital capacity (FVC) ratio [1]. Previously, this pattern called restricted pulmonary function had not been classified, as it did not meet the diagnostic criteria for restrictive nor obstructive ventilatory impairment commonly used to defined chronic obstructive pulmonary disease (COPD)[2–4]. However, recent epidemiological studies, mainly conducted in Europe and North America, have shown its high prevalence among smokers, and in the general population. In smokers, PRISm has been shown to be associated with a high burden of respiratory morbidity and mortality [5, 6]. In the general population, a similar relationship has been demonstrated between PRISm, poor cardiovascular diseases (CVD), and all-cause mortality outcomes [7–15].

To date, there has only been few similar studies conducted in Asian populations (i.e., Japan, Korea), which were limited by the small sample size and cross-sectional study design [8, 9, 16, 17]. Moreover, the limited evidence that is available, suggest that the prevalence of PRISm across different populations may be variable, ranging from 4% to 48%[1, 10, 12, 14, 18]. This may in part relate to differences in population demographics as PRISm has been shown to be more common in older adults, smokers and in comorbid conditions such as asthma, and metabolic abnormalities [16, 19]. Furthermore, differences in the reference values used to interpret spirometric data between populations especially from different ethnic background, may also contribute to the variation in reported rates of PRISm [10, 20]. Therefore, the evidence published thus far, may not be applicable to the Chinese population. The objectives of the present study were to estimate the prevalence of PRISm in the general Chinese population using data from the Prospective Urban Rural Epidemiology (PURE) study; and to examine for the associations between PRISm with future all-cause mortality, and poor CVD outcomes.

## Methods

### Study design and participants

This report includes 47,677 participants enrolled from 115 communities (70 urban and 45 rural) across 12 provinces in China as part of the PURE study (**Figure S1**) [21]. Participants were recruited via three-level cluster sampling method (province, community, and household) between 2005-2009, and followed up until August 11, 2022. Details of study design and sampling are described in online supplementary **Appendix A1**. The study protocol was approved by the ethics committees of the Fuwai Hospital of Chinese Academy of Medical Sciences and Beijing Hypertension League Institute. Informed consent was obtained from all participants prior to data collection. This study was reported in accordance with the STROBE guideline [22].

### Baseline data collection

The questionnaire collected data on participants’ demographic and medical information, including physician-diagnosed medical conditions, symptoms, and regular medication use. Demographic data included socioeconomic status (SES), such as education, occupation, and household wealth index; lifestyle behaviors (tobacco and alcohol use, second-hand smoking, dietary pattern and physical activity); and air pollution exposures (particulate matter (PM) 2.5, and household cooking fuels). Physical measurements included body mass index (BMI), waist-hip ratio (WHR), and handgrip strength. More details in **Appendix A2 and Table S1**.

Anthropometric parameters (weight, height, waist and hip circumferences) were measured using a standardized protocol. Blood samples were collected according to the relevant guidelines in the Third Xiangya Hospital. Fasting blood samples were collected to test total cholesterol (TC), and triglycerides (TG) using LEADMAN test kits (Beijing LEADMAN Biochemical Co., Ltd. China) [23].

### Lung function measurements

Lung function was measured with a portable spirometer (MicroGP; MicroMedical, Chatham, IL, USA), without spirographs, using a standardized protocol, following the 2005 American Thoracic Society and European Respiratory Society (ATS/ERS) guidelines [24]. Participants were coached before attempting pre-bronchodilator forced expiratory maneuvers (maximum six attempts) while standing and wearing a nose-clip, although the recent 2022 ATS/ERS guidelines using post-bronchodilator spirometry [3]. Maneuvers were observed for maximal effort, forced exhalation time >6 seconds, and exhalation without coughing. Spirometers were calibrated monthly (3L syringe) or before each use in extreme temperature or handling. For analyses, we selected participants with at least two acceptable FEV_1_ and FVC measurements within 200 mL variability. The quality of the spirometry data in PURE have previously been validated for external [20], and face validity [25](**Appendix A3)**. The highest FEV_1_, FVC, and their ratio recorded for individual were used in this analysis.

Participants were categorized into 1 of 3 groups according to their baseline pre-bronchodilator spirometry finding in this study. The Global Lung Initiative (GLI) reference values appropriate for age, sex, and height for North East Asian were used to interpret the spirometry data [26]. Participants with a FEV_1_<80% predicted and FEV_1_/FVC ≥0.70 were categorized into the PRISm group; Airflow obstruction (AO) included participants with a FEV_1_/FVC<0.70; while a FEV_1_ ≥80% predicted and a FEV_1_/FVC ratio ≥ 0.70 were considered as normal [16, 27].

### Assessment of outcomes

Follow-up was conducted every three years to obtain information on major cardiovascular events and mortality (classified by causes) using standardized case report forms. All deaths and major cardiovascular events were adjudicated centrally in Beijing by the trained Clinical Event Committee (CEC) using standard definitions and the International Classification of Diseases (ICD)-10 code according to a standard protocol as previously described [28]. The outcomes of this study were all-cause mortality, CVD mortality, major CVD (composite of non-fatal myocardial infarction [MI], stroke, and heart failure [HF]) and its components, as well as respiratory mortality (**Appendix A4)**.

### Statistical analysis

The baseline characteristics and lung function data according to spirometry subgroups (i.e., PRISm, AO, Normal) were summarized using descriptive statistics. Categorical variables are presented as counts and proportions, normally distributed variables as means and standard deviations (SD); and non-normally distributed variables as median and interquartile range (IQR). Crude death rates and incidences of CVD events are reported as percentages and 1000 person-years. Kaplan–Meier survival analysis was used to determine the association between lung function with follow up clinical outcomes. Cox proportional hazard regression was used to evaluate the relationship between lung function groups and outcomes, adjusting for covariates with centre as a random effect variable (to account for clustering sampling). Proportionality was checked using Schoenfeld residuals [29]. For outcome assessed, participants were censored either at the time they were lost to follow-up (if they could not be contacted or requested to withdraw from the study), or at the date of the last follow-up. The base model was adjusted for age, sex and location(rural/urban). The fully adjusted model was further adjusted for education, occupation, household wealth index, tobacco consumption, pack-year, second-hand smoking exposure, alcohol consumption, BMI, WHR, physical activity, handgrip strength, household cooking fuel, PM 2.5, serum TC, TG, comorbidities (COPD, asthma, CVDs, hypertension, diabetes) and regular medications (i.e. antihypertensive drugs, cholesterol lowering drug, asthma drug and others). Subgroup analyses were performed by lung function groups. Potential multiplicative interaction was explored between PRISm and socio-demographic/lifestyle factors on the primary outcomes, including age, sex, location, smoking status (current and former smokers vs non-smokers), overweight, education level, and PM 2.5 exposure.

Several sensitivity analyses were conducted to validate the robustness of the principal findings, including: (1) Defining the lung function groups by the Lower Limits of Normal (LLN) criteria; (2) Participants without self-reported COPD or asthma at baseline; (3) Participants without pre-existing CVDs at baseline; (4) Excluding participants who developed clinical outcomes within the first 2 years of follow-up to minimize the risk of reverse causality; (5) Excluding participants in the lowest quartile of handgrip strength to minimize the confounding of severe weakness or frailty on low pulmonary function tests; (6) Additional adjustment for differences in the severity of FEV_1_ impairment across groups (as a continuous variable); (7) Additional adjustment for respiratory exacerbations.

We assessed the adequacy of the sample size using guidelines proposed by Concato and colleagues [30], which recommends that for Cox regression at least ten events for each degree of freedom (df) are needed to provide stable models. The number of events during follow-up was greater than ten events per df. Missing data imputation were not conducted in this study.

All analyses and graphs were generated using SAS software, version 9.4 (SAS Institute Inc., Cary, North Carolina, USA) and R software, version 4.2.1 (R Foundation for Statistical Computing).

## Results

Of the 47,677 participants enrolled from 70 urban and 45 rural communities across 12 provinces in China, 45,889 had complete baseline data and spirometry measurements (**Figure 1**). Of these, 4,878 were excluded due to unacceptable FEV_1_ or FVC and 732 had missing smoking data, leaving 40,279 participants (84.4%) in the final analysis.

**Figure 1.**
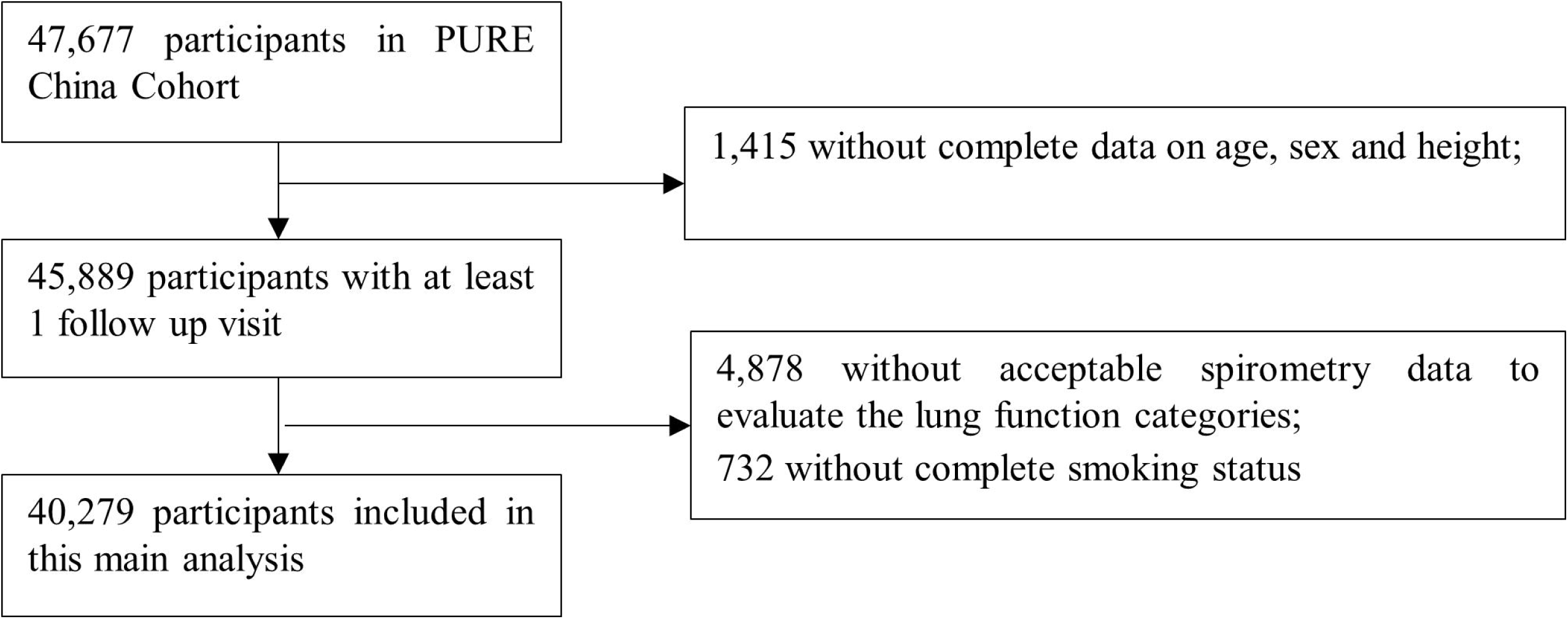
Flow diagram of PURE-China Study for this analysis.

**Table 1** summarizes the baseline characteristics for the three spirometry subgroups. There were 11,824 (29.3%) participants with baseline spirometry finding for PRISm, 3,407 (8.5%) for AO, and 25,048 (62.2%) with normal spirometry. Compared to the normal group, participants with PRISm were on average slightly younger (50.7 vs 51 yrs), and there were higher proportion of males (43% vs 39.9%), urban residents (52.6% vs 50%), smokers (27.8% vs 26.3%), and comorbidities including morbid obesity (44.8% vs 42.5%). Participants with AO were generally older (53.6 yrs) with lower SES (43.2%), smokers (greater intensity of smoking, 30.3% with 23 pack-years), and high burden of comorbidities compared to the normal group. The overall smoking rate (either current or former smoker) was 27.1%, with higher rates observed in the AO (30.3%) and PRISm (27.8%) groups.

**Table 1:**
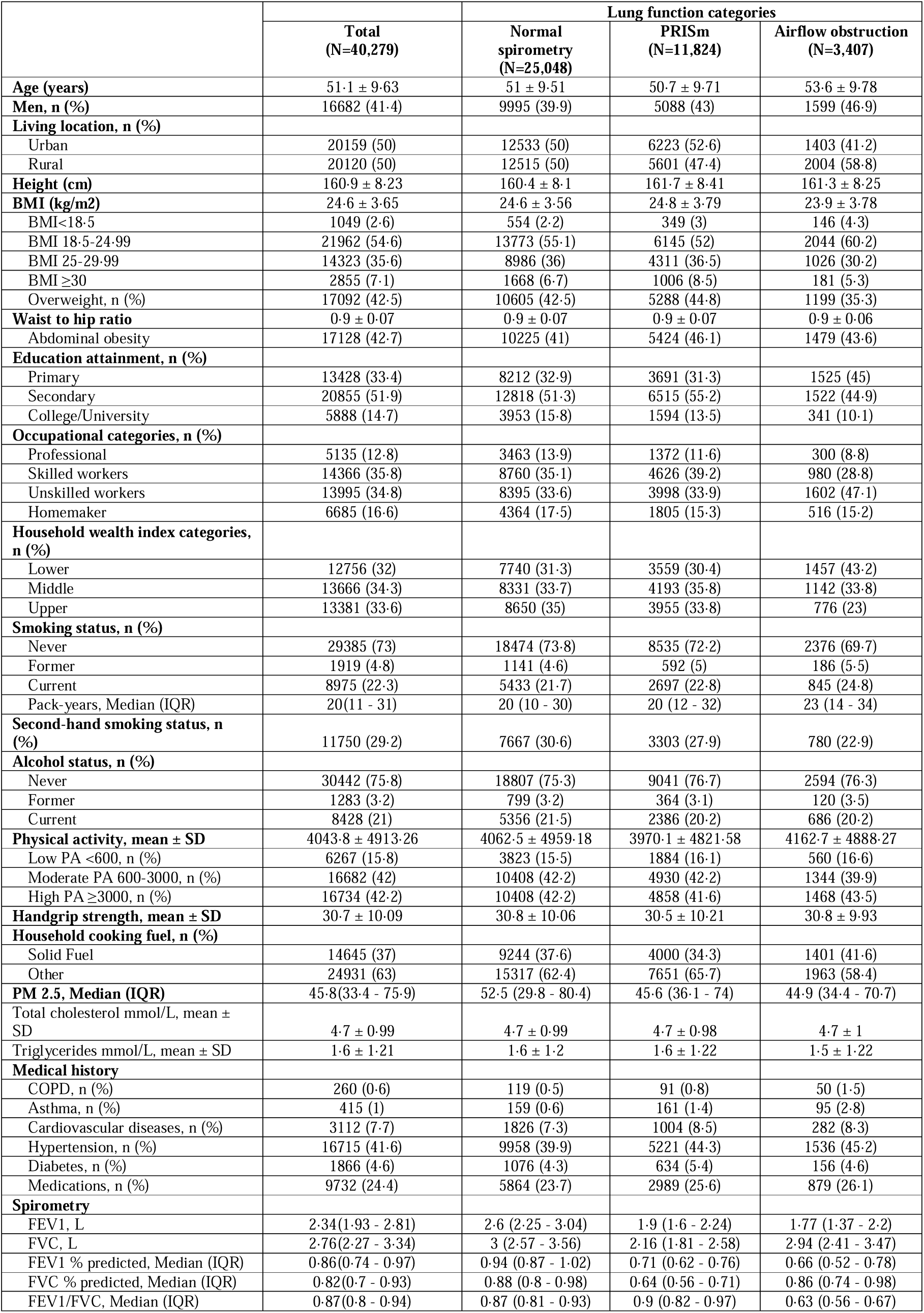
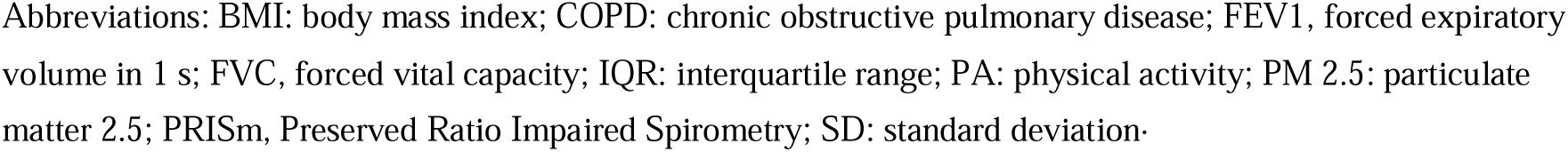
Baseline characteristics of the overall study population according to the baseline lung function categories.

|  | Total<br>(N=40,279) | Lung function categories |  |  |
| --- | --- | --- | --- | --- |
|  |  | Normal<br>spirometry<br>(N=25,048) | PRISm<br>(N=11,824) | Airflow obstruction<br>(N=3,407) |
| Age (years) | 51.1 ± 9.63 | 51 ± 9.51 | 50.7 ± 9.71 | 53.6 ± 9.78 |
| Men, n (%) | 16682 (41.4) | 9995 (39.9) | 5088 (43) | 1599 (46.9) |
| Living location, n (%) |  |  |  |  |
| Urban | 20159 (50) | 12533 (50) | 6223 (52.6) | 1403 (41.2) |
| Rural | 20120 (50) | 12515 (50) | 5601 (47.4) | 2004 (58.8) |
| Height (cm) | 160.9 ± 8.23 | 160.4 ± 8.1 | 161.7 ± 8.41 | 161.3 ± 8.25 |
| BMI (kg/m<sup>2</sup>) | 24.6 ± 3.65 | 24.6 ± 3.56 | 24.8 ± 3.79 | 23.9 ± 3.78 |
| BMI < 18.5 | 1049 (2.6) | 554 (2.2) | 349 (3) | 146 (4.3) |
| BMI 18.5-24.99 | 21962 (54.6) | 13773 (55.1) | 6145 (52) | 2044 (60.2) |
| BMI 25-29.99 | 14323 (35.6) | 8986 (36) | 4311 (36.5) | 1026 (30.2) |
| BMI ≥ 30 | 2855 (7.1) | 1668 (6.7) | 1006 (8.5) | 181 (5.3) |
| Overweight, n (%) | 17092 (42.5) | 10605 (42.5) | 5288 (44.8) | 1199 (35.3) |
| Waist to hip ratio | 0.9 ± 0.07 | 0.9 ± 0.07 | 0.9 ± 0.07 | 0.9 ± 0.06 |
| Abdominal obesity | 17128 (42.7) | 10225 (41) | 5424 (46.1) | 1479 (43.6) |
| Education attainment, n (%) |  |  |  |  |
| Primary | 13428 (33.4) | 8212 (32.9) | 3691 (31.3) | 1525 (45) |
| Secondary | 20855 (51.9) | 12818 (51.3) | 6515 (55.2) | 1522 (44.9) |
| College/University | 5888 (14.7) | 3953 (15.8) | 1594 (13.5) | 341 (10.1) |
| Occupational categories, n (%) |  |  |  |  |
| Professional | 5135 (12.8) | 3463 (13.9) | 1372 (11.6) | 300 (8.8) |
| Skilled workers | 14366 (35.8) | 8760 (35.1) | 4626 (39.2) | 980 (28.8) |
| Unskilled workers | 13995 (34.8) | 8395 (33.6) | 3998 (33.9) | 1602 (47.1) |
| Homemaker | 6685 (16.6) | 4364 (17.5) | 1805 (15.3) | 516 (15.2) |
| Household wealth index categories, n (%) |  |  |  |  |
| Lower | 12756 (32) | 7740 (31.3) | 3559 (30.4) | 1457 (43.2) |
| Middle | 13666 (34.3) | 8331 (33.7) | 4193 (35.8) | 1142 (33.8) |
| Upper | 13381 (33.6) | 8650 (35) | 3955 (33.8) | 776 (23) |
| Smoking status, n (%) |  |  |  |  |
| Never | 29385 (73) | 18474 (73.8) | 8535 (72.2) | 2376 (69.7) |
| Former | 1919 (4.8) | 1141 (4.6) | 592 (5) | 186 (5.5) |
| Current | 8975 (22.3) | 5433 (21.7) | 2697 (22.8) | 845 (24.8) |
| Pack-years, Median (IQR) | 20(11 - 31) | 20 (10 - 30) | 20 (12 - 32) | 23 (14 - 34) |
| Second-hand smoking status, n (%) | 11750 (29.2) | 7667 (30.6) | 3303 (27.9) | 780 (22.9) |
| Alcohol status, n (%) |  |  |  |  |
| Never | 30442 (75.8) | 18807 (75.3) | 9041 (76.7) | 2594 (76.3) |
| Former | 1283 (3.2) | 799 (3.2) | 364 (3.1) | 120 (3.5) |
| Current | 8428 (21) | 5356 (21.5) | 2386 (20.2) | 686 (20.2) |
| Physical activity, mean ± SD | 4043.8 ± 4913.26 | 4062.5 ± 4959.18 | 3970.1 ± 4821.58 | 4162.7 ± 4888.27 |
| Low PA < 600, n (%) | 6267 (15.8) | 3823 (15.5) | 1884 (16.1) | 560 (16.6) |
| Moderate PA 600-3000, n (%) | 16682 (42) | 10408 (42.2) | 4930 (42.2) | 1344 (39.9) |
| High PA ≥ 3000, n (%) | 16734 (42.2) | 10408 (42.2) | 4858 (41.6) | 1468 (43.5) |
| Handgrip strength, mean ± SD | 30.7 ± 10.09 | 30.8 ± 10.06 | 30.5 ± 10.21 | 30.8 ± 9.93 |
| Household cooking fuel, n (%) |  |  |  |  |
| Solid Fuel | 14645 (37) | 9244 (37.6) | 4000 (34.3) | 1401 (41.6) |
| Other | 24931 (63) | 15317 (62.4) | 7651 (65.7) | 1963 (58.4) |
| PM 2.5, Median (IQR) | 45.8(33.4 - 75.9) | 52.5 (29.8 - 80.4) | 45.6 (36.1 - 74) | 44.9 (34.4 - 70.7) |
| Total cholesterol mmol/L, mean ± SD | 4.7 ± 0.99 | 4.7 ± 0.99 | 4.7 ± 0.98 | 4.7 ± 1 |
| Triglycerides mmol/L, mean ± SD | 1.6 ± 1.21 | 1.6 ± 1.2 | 1.6 ± 1.22 | 1.5 ± 1.22 |
| Medical history |  |  |  |  |
| COPD, n (%) | 260 (0.6) | 119 (0.5) | 91 (0.8) | 50 (1.5) |
| Asthma, n (%) | 415 (1) | 159 (0.6) | 161 (1.4) | 95 (2.8) |
| Cardiovascular diseases, n (%) | 3112 (7.7) | 1826 (7.3) | 1004 (8.5) | 282 (8.3) |
| Hypertension, n (%) | 16715 (41.6) | 9958 (39.9) | 5221 (44.3) | 1536 (45.2) |
| Diabetes, n (%) | 1866 (4.6) | 1076 (4.3) | 634 (5.4) | 156 (4.6) |
| Medications, n (%) | 9732 (24.4) | 5864 (23.7) | 2989 (25.6) | 879 (26.1) |
| Spirometry |  |  |  |  |
| FEV1, L | 2.34(1.93 - 2.81) | 2.6 (2.25 - 3.04) | 1.9 (1.6 - 2.24) | 1.77 (1.37 - 2.2) |
| FVC, L | 2.76(2.27 - 3.34) | 3 (2.57 - 3.56) | 2.16 (1.81 - 2.58) | 2.94 (2.41 - 3.47) |
| FEV1 % predicted, Median (IQR) | 0.86(0.74 - 0.97) | 0.94 (0.87 - 1.02) | 0.71 (0.62 - 0.76) | 0.66 (0.52 - 0.78) |
| FVC % predicted, Median (IQR) | 0.82(0.7 - 0.93) | 0.88 (0.8 - 0.98) | 0.64 (0.56 - 0.71) | 0.86 (0.74 - 0.98) |
| FEV1/FVC, Median (IQR) | 0.87(0.8 - 0.94) | 0.87 (0.81 - 0.93) | 0.9 (0.82 - 0.97) | 0.63 (0.56 - 0.67) |

Abbreviations: BMI: body mass index; COPD: chronic obstructive pulmonary disease; FEV1, forced expiratory volume in 1 s; FVC, forced vital capacity; IQR: interquartile range; PA: physical activity; PM 2.5: particulate matter 2.5; PRISm, Preserved Ratio Impaired Spirometry; SD: standard deviation

During a median follow-up of 11.9 (IQR 9.5-12.5) years (no significant difference among the three groups), there were 2,214 (5.5%) deaths, 773 (1.9%) CVD deaths, and 3,507 (8.71%) major CVD events including 939 (2.3%) MI, 2,483 (6.2%) strokes, 277 (0.7%) HF and 77 respiratory deaths. The crude cumulative rates of participants without clinical events during follow-up were significantly lower for PRISm and AO compared to normal (**Figure 2**). **Table 2** highlight the associations between baseline spirometry groups with future clinical events. In the fully adjusted model 2, PRISm remained significantly associated with all-cause mortality (HR=1.42 [95%CI: 1.29-1.58]), CVD mortality (HR=1.35 [1.14-1.60]), major CVD events (HR=1.16 [1.07-1.25]), MI (HR=1.34 [1.15-1.56]), and HF (HR=2.02 [1.46-2.79]); but not with strokes (HR=0.98 [0.85-1.13]). Similarly, the fully adjusted model 2, showed significant associations between AO with all-cause mortality, CVD mortality, and MI; but not HF, major CVD or strokes. In addition, the effect estimates comparing PRISm to AO were presented in **Table S2,** indicating that higher risk pattern was observed among PRISm participants.

**Figure 2.**
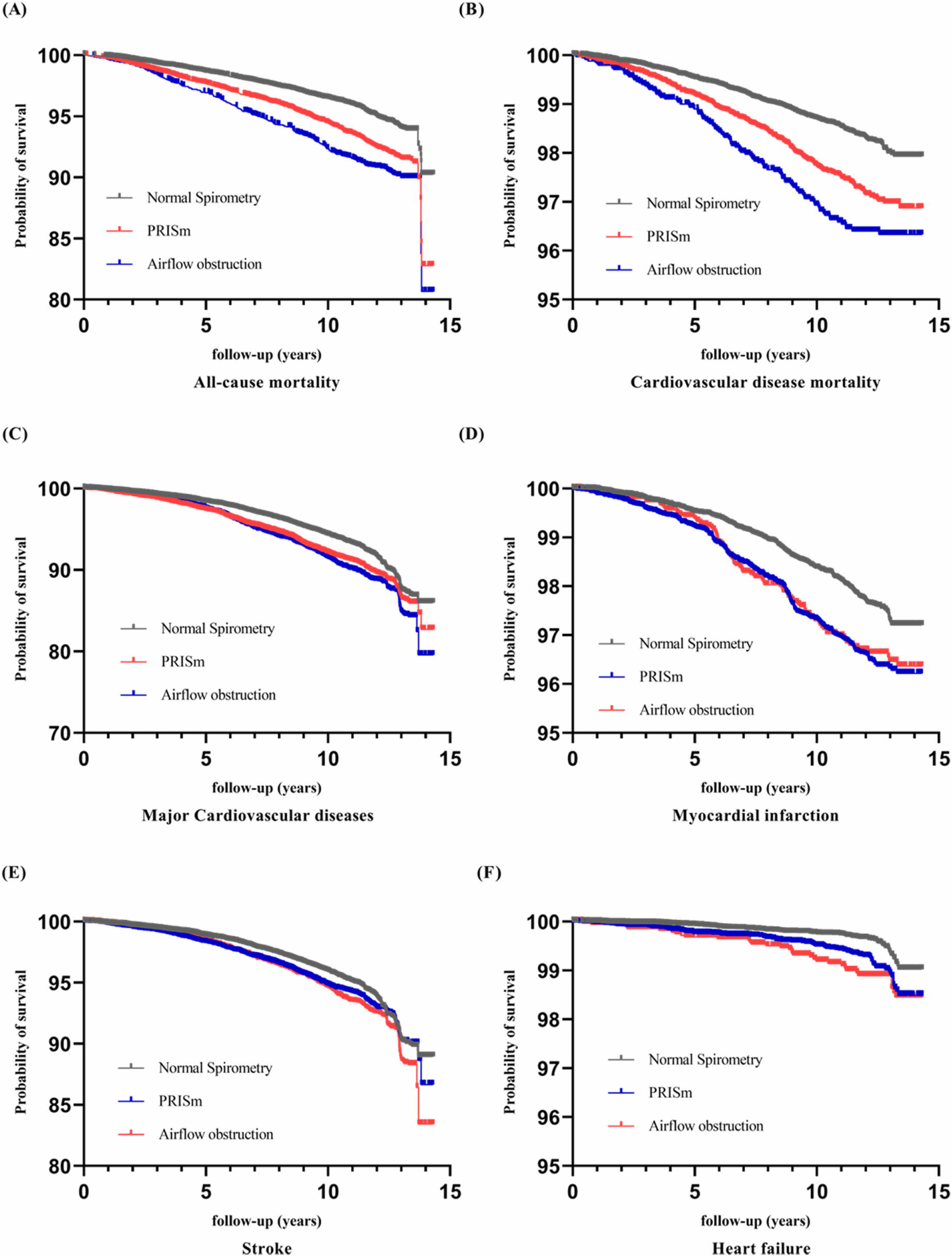
Kaplan-Meier curve by baseline Lung Function categories.

**Table 2:**
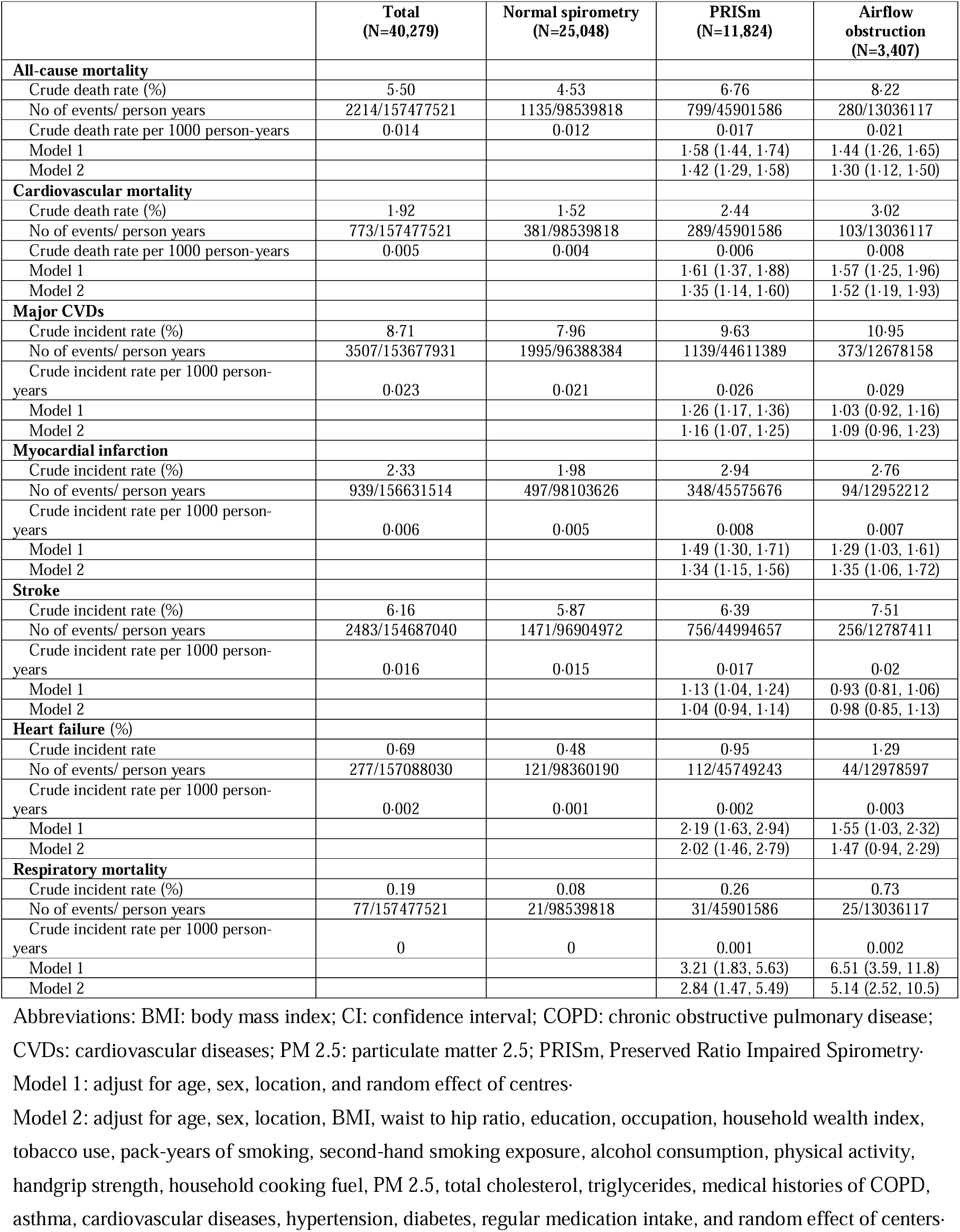
Hazard ratios (95%CI) of different outcomes with lung function categories.

|  | Total<br>(N=40,279) | Normal spirometry<br>(N=25,048) | PRISm<br>(N=11,824) | Airflow<br>obstruction<br>(N=3,407) |
| --- | --- | --- | --- | --- |
| **All-cause mortality** |  |  |  |  |
| Crude death rate (%) | 5.50 | 4.53 | 6.76 | 8.22 |
| No of events/ person years | 2214/157477521 | 1135/98539818 | 799/45901586 | 280/13036117 |
| Crude death rate per 1000 person-years | 0.014 | 0.012 | 0.017 | 0.021 |
| Model 1 |  |  | 1.58 (1.44, 1.74) | 1.44 (1.26, 1.65) |
| Model 2 |  |  | 1.42 (1.29, 1.58) | 1.30 (1.12, 1.50) |
| **Cardiovascular mortality** |  |  |  |  |
| Crude death rate (%) | 1.92 | 1.52 | 2.44 | 3.02 |
| No of events/ person years | 773/157477521 | 381/98539818 | 289/45901586 | 103/13036117 |
| Crude death rate per 1000 person-years | 0.005 | 0.004 | 0.006 | 0.008 |
| Model 1 |  |  | 1.61 (1.37, 1.88) | 1.57 (1.25, 1.96) |
| Model 2 |  |  | 1.35 (1.14, 1.60) | 1.52 (1.19, 1.93) |
| **Major CVDs** |  |  |  |  |
| Crude incident rate (%) | 8.71 | 7.96 | 9.63 | 10.95 |
| No of events/ person years | 3507/153677931 | 1995/96388384 | 1139/44611389 | 373/12678158 |
| Crude incident rate per 1000 person-years | 0.023 | 0.021 | 0.026 | 0.029 |
| Model 1 |  |  | 1.26 (1.17, 1.36) | 1.03 (0.92, 1.16) |
| Model 2 |  |  | 1.16 (1.07, 1.25) | 1.09 (0.96, 1.23) |
| **Myocardial infarction** |  |  |  |  |
| Crude incident rate (%) | 2.33 | 1.98 | 2.94 | 2.76 |
| No of events/ person years | 939/156631514 | 497/98103626 | 348/45575676 | 94/12952212 |
| Crude incident rate per 1000 person-years | 0.006 | 0.005 | 0.008 | 0.007 |
| Model 1 |  |  | 1.49 (1.30, 1.71) | 1.29 (1.03, 1.61) |
| Model 2 |  |  | 1.34 (1.15, 1.56) | 1.35 (1.06, 1.72) |
| **Stroke** |  |  |  |  |
| Crude incident rate (%) | 6.16 | 5.87 | 6.39 | 7.51 |
| No of events/ person years | 2483/154687040 | 1471/96904972 | 756/44994657 | 256/12787411 |
| Crude incident rate per 1000 person-years | 0.016 | 0.015 | 0.017 | 0.02 |
| Model 1 |  |  | 1.13 (1.04, 1.24) | 0.93 (0.81, 1.06) |
| Model 2 |  |  | 1.04 (0.94, 1.14) | 0.98 (0.85, 1.13) |
| **Heart failure (%)** |  |  |  |  |
| Crude incident rate | 0.69 | 0.48 | 0.95 | 1.29 |
| No of events/ person years | 277/157088030 | 121/98360190 | 112/45749243 | 44/12978597 |
| Crude incident rate per 1000 person-years | 0.002 | 0.001 | 0.002 | 0.003 |
| Model 1 |  |  | 2.19 (1.63, 2.94) | 1.55 (1.03, 2.32) |
| Model 2 |  |  | 2.02 (1.46, 2.79) | 1.47 (0.94, 2.29) |
| **Respiratory mortality** |  |  |  |  |
| Crude incident rate (%) | 0.19 | 0.08 | 0.26 | 0.73 |
| No of events/ person years | 77/157477521 | 21/98539818 | 31/45901586 | 25/13036117 |
| Crude incident rate per 1000 person-years | 0 | 0 | 0.001 | 0.002 |
| Model 1 |  |  | 3.21 (1.83, 5.63) | 6.51 (3.59, 11.8) |
| Model 2 |  |  | 2.84 (1.47, 5.49) | 5.14 (2.52, 10.5) |

Abbreviations: BMI: body mass index; CI: confidence interval; COPD: chronic obstructive pulmonary disease; CVDs: cardiovascular diseases; PM 2.5: particulate matter 2.5; PRISm, Preserved Ratio Impaired Spirometry. Model 1: adjust for age, sex, location, and random effect of centres. Model 2: adjust for age, sex, location, BMI, waist to hip ratio, education, occupation, household wealth index, tobacco use, pack-years of smoking, second-hand smoking exposure, alcohol consumption, physical activity, handgrip strength, household cooking fuel, PM 2.5, total cholesterol, triglycerides, medical histories of COPD, asthma, cardiovascular diseases, hypertension, diabetes, regular medication intake, and random effect of centers.

**Table 3** presents the associations between PRISm with different outcomes stratified by smoking status compared to normal spirometry. The findings indicate a similar direction of association between PRISm in current or former smokers and non-smokers with all-cause mortality, major CVD, and HF. But the association is a little greater among non-smokers vs smokers, especially for CVD mortality.

**Table 3:** Hazard ratios (95%CI) for different outcomes with PRISm in smokers and non-smokers.

| Outcomes | PRISm among smokers (N=3289) | PRISm among non-smokers (N=8535) |
| --- | --- | --- |
| All-cause mortality | 1.42 (1.21, 1.65) | 1.43 (1.25, 1.63) |
| Cardiovascular mortality | 1.14 (0.87, 1.48) | 1.52 (1.22, 1.89) |
| Major CVDs | 1.23 (1.08, 1.41) | 1.11 (1.01, 1.23) |
| Myocardial infarction | 1.25 (0.98, 1.60) | 1.40 (1.16, 1.69) |
| Stroke | 1.15 (0.98, 1.35) | 0.98 (0.87, 1.11) |
| Heart failure | 2.35 (1.33, 4.16) | 1.84 (1.25, 2.72) |
| Respiratory mortality | 3.38 (1.16, 9.88) | 2.28 (0.68, 7.70) |

Abbreviations: BMI, body mass index; CVD, cardiovascular disease; FEV1, forced expiratory volume in 1 s; FVC, forced vital capacity; LLN, lower limit normal; PM 2.5: particulate matter 2.5; PRISm, Preserved Ratio Impaired Spirometry. Full model adjusted for age, sex, location, BMI, waist to hip ratio, education, occupation, household wealth index, tobacco use, pack-years of smoking, second-hand smoking exposure, alcohol consumption, physical activity, handgrip strength, household cooking fuel, PM 2.5, total cholesterol, triglycerides, medical histories of COPD, asthma, cardiovascular diseases, hypertension, diabetes, regular medication intake, and random effect of centers in this analysis. Individuals with normal spirometry were used as the reference group.

Other stratified analyses shown in **Figure S2,** suggested higher risks for PRISm (compared to normal spirometry) with all-cause mortality in rural communities versus urban (HR=1.55 [1.37-1.76] *vs* HR=1.28 [1.07-1.53]; p-interaction 0.044); and normal versus high BMI (HR=1.50 [1.31-1.72] *vs* HR=1.38 [1.18-1.61]; p-interaction 0.042). Other analyses stratified by age, sex, smoking status, PM2.5 exposure level, and educational level, showed no difference in associations between PRISm with clinical outcomes. Nevertheless, all stratified analyses should be considered as exploratory due to multiple testing.

A number of different sensitivity analyses were conducted to assess the robustness of the association (**Table S3**). These include excluding participants with self-reported COPD, asthma, CVD, and the lowest quartile of handgrip strength, and those with incident CVD events in the first two years of follow-up. There were no material differences found in the direction or magnitude of risk with PRISm on each of the clinical outcome. Similar findings when using the LLN thresholds to categorize the spirometry groups. Lastly, FEV_1_ as a continuous variable was added into the model to account for any differences in the severity of lung function impairment across groups. While, the HR for PRISm on all-cause mortality, major CVD, MI, and HF showed a slight decrease in magnitude, these remained statistically significant, except for CVD mortality.

## Discussion

In this nationwide, community-based Chinese population, the prevalence of PRISm on spirometry was high, at 29.3% of the study population. PRISm was associated with significantly higher risk for all-cause mortality, CVD mortality, future MI, and HF. The relationship was consistent across different subgroups stratified by smoking status, demographic and socio-economic factors. To the best of our knowledge, this is the first prospective study to examine the relationship between PRISm with all-cause mortality and CVD outcomes within a broad Chinese population.

Previous large-scale population-based studies conducted in Europe, the United States, Japan, and South Korea, have reported variable prevalence of PRISm on spirometry in the general population ranging from 6.2% to 16.7% [1, 7, 9, 11, 14, 31]. In two other studies conducted in older adults and in hospital-based centres, the prevalence of PRISm exceeded 20% [12, 13]. Subgroup analysis of the Burden of Lung Disease study has shown that the prevalence of PRISm in Guangzhou, China was 23.4%, higher than the global average of 11.7% [18]. This is similar to our estimate of 29.3% in a community-based cohort sampled from urban and rural communities across 12 provinces in China. High tobacco smoke exposure (either from smoking, or second-hand exposure) may contribute to the high rates of PRISm in the Chinese population, as cigarette smoking has been shown to be an important risk factor for both PRISm and COPD [10, 17, 32, 33]. While the rates of cigarette smoking have declined in many developed countries over the past thirty years, cigarette consumption in China has substantially increased [34]. Consequently, the reported prevalence of COPD in the Chinese population (8.6% in the general population and 13.7% for those over 40 years of age) exceeds the global average of 3.9% [19, 32]. Furthermore, ambient exposure to PM2.5, has been linked to the development of COPD [35, 36], and whether this is also a risk factor for PRISm requires further investigation. There is a strong reason to suspect that the higher incidence of PRISm in the Chinese population may in part be related to air pollution in China, since others suggested that air pollution and organic inhalational exposures might cause PRISm through systemic inflammation in middle-income countries[37]. In any case, public health strategies which reduces tobacco smoking and air pollution can have substantial impact in mitigating the high prevalence of PRISm and burden of adverse CVD outcomes in the Chinese population.

Our main analysis clearly showed that baseline PRISm was associated with significantly higher risks for long-term all-cause mortality, CVD mortality, major CVD events, including non-fatal MI, and HF. The findings were robust and consistent across different sensitivity analyses; including using the LLN approach to reclassify PRISm, and excluding participants with established CVD, COPD or asthma, to avoid reverse causality. Additionally, the fact that adjusting for baseline FVC in the comprehensive model indicates that the negative health impacts of PRISm occur independently of FVC. Furthermore, our findings are concordant with other studies conducted in Europe, America, and Japan [7, 9–11, 13, 14], thus supporting the generalizability of these associations. The one exception was in a study in Korea, which showed no correlation between PRISm with all-cause mortality and CVD mortality[8]. In addition, we found that PRISm was associated with an elevated risk of all-cause mortality by 42% and CVD mortality by 35%; which are lower than previously reported estimates between 50%-120% for all-cause mortality [9–11, 13, 14] and 55%-307% for CVD mortality [7, 9, 11, 13, 14]. Only one other longitudinal study has examined the association of PRISm with MI, stroke, and HF as separate endpoints, and showed a significant positive correlation with these CVD outcomes[7]. Consistent with these findings, we also found a positive association between PRISm with MI and HF, but not with strokes.

Another noteworthy finding was a trend for higher risk of poor long-term outcome with PRISm than for AO, even though the differences were not statistically significant. Similar findings have been reported by the Rotterdam Study, NHLBI Study, and UK Biobank Study [7, 11, 14]. It is important to note that the current spirometry definition for PRISm may include a heterogeneous group of conditions ranging from obstructive diseases with smaller airways obstruction to restrictive lung pathology. In fact, a recent study had reported on several different phenotypes within PRISm[1], which were identified as either COPD-, Restrictive-, or Metabolic-subtypes. Indeed, research utilizing computerized tomography (CT) of the lungs have shown distinctive characteristics, which distinguished PRISm from normal individuals. These include findings of inflammation and remodeling in the respiratory bronchioles and peripheral alveolar tissue, resulting in functional small airway disease (fSAD), and reduction in the pulmonary small vasculature in PRISm versus normal individuals [38]. Other studies have suggested that the main differences between PRISm and COPD may be lower burden of parenchymal damage and emphysematous changes [39]. More work is needed to inform the underlying pathophysiology and pathogenesis of PRISm, which may help us to better understand why PRISm is associated with worse health outcomes.

We also uncovered a number of novel and interesting findings from our stratified analyses. First, the associations between PRISm with poor outcomes were similar in both smokers and non-smokers, even greater in non-smokers, indicating equally worse outcome in patients with PRISm associated with non-smoking etiological factors. This finding aligns with some previous stratified studies, indicating that PRISm increased all-cause death risks in both smokers and non-smokers [9, 13], with some studies suggesting an even higher risk among the non-smoker [11, 27]. It also suggested that smoking is unlikely to be a confounding factor for the association between PRISm with CVD events. Second, we found that PRISm was associated with a worse outcome in rural compared to urban communities. This may relate to the lower SES and related factors such as poor access to medical resources and adverse early life conditions, which are important determinants of health but were not well captured in this study. Third, participants with normal BMI did worse than those with elevated BMI with PRISm, which is consistent with Wan’s finding [11] but contrary to Higbee’s analyses [10]. We speculate, on the one hand, it might be related to the finding of PRISm in obese individuals that was representative of reduced chest compliance leading to impaired ventilatory capacity [40], rather than any substantial pathology in the airways or lung parenchyma. On the other hand, there has been an association between COPD and a low body weight, referred to as the "obesity paradox" [41]. Data from Asian countries showed that COPD patients with high BMI tend to have less symptom, better health-related quality of life and lung function, and fewer exacerbation and death [42]. The remaining stratified analyses, by age, sex, PM 2.5 exposure and education level, suggest that the health risks attributable to PRISm were not modified by these factors.

The strengths of this study include the prospective long-term follow-up, the high-quality and standardized methodology in data collection and the large sample size, which allows for comprehensive adjustments of the potential confounding factors, increase study power and validity to our findings. Secondly, we performed multiple sensitivity and stratified analyses to ensure the robustness and authenticity of our findings.

This study has several limitations. Firstly, according to the Global Initiative for Chronic Obstructive Lung Disease 2023 Report[43], spirometry tests and classification (including PRISm) should be performed post-bronchodilator inhalation. However, in this study, all spirometry assessments were conducted pre-bronchodilator, which may lead to a potential misclassification of some patients with reversible FEV1 reduction as PRISm, thereby potentially overestimating the prevalence of PRISm. Another similar concern is that lung function in this study was measured using a spirometer without spirographs. Despite our best efforts to ensure the quality of pulmonary function test data (**Appendix A3**), there still might be instances of underestimated FVC and artificially normalized FEV1/FVC ratios due to premature termination of exhalation. This suggests that future studies should aim to optimize the accuracy of pulmonary function testing to obtain more precise correlations between PRISm and clinical outcomes. However, it should be noted that utilizing spirometry with spirographs and conducting post-bronchodilator tests would reduce feasibility in community population screenings. Additionally, due to the internal heterogeneity of PRISm, patients identified with PRISm still require further precise examinations after diagnosis, rather than direct targeted treatment interventions. Therefore, we believe that PRISm, as a previously overlooked pulmonary function phenotype, holds significant screening value in the field of public health and warrants increased attention. Furthermore, other lung function parameters such as lung volumes or diffusion capacity were not collected, thus limiting our ability to examine the heterogeneity in phenotypes within PRISm. Lastly, as this is an observational study, any inferences on a causality must be made cautiously, and further trajectory of PRISm research is needed to explore whether this association is causal.

## Conclusion

In conclusion, our findings highlight the high prevalence of PRISm in the general Chinese population, and support prior reports of the poor CVD outcomes with PRISm in other diverse populations. The early prevention, diagnosis, and intervention of PRISm may offer an important strategy to alleviating the high CVD burden in China. However, further studies are needed to improve our current understanding of the pathophysiology of PRISm, and unraveling the mechanisms underlying its strong relationship with poor CVD outcomes.

## Data Availability

The datasets analysed during the current study are not publicly available due to ongoing project, but are available from the corresponding author on reasonable request.

## Author Contributors

**Conceptualization:** Mengya Li, Yang Li, Duong Mylinh, Lap Ah Tse, Zhiguang Liu

**Data curation:** Sumathy Rangarajan

**Formal analysis:** Mengya Li

**Funding acquisition:** Salim Yusuf, Wei Li, Bo Hu

**Investigation:** Deren Qiang, Kai You, Jiying Li, Bo Hu

**Methodology:** Mengya Li, Yang Li, Zhiguang Liu

**Project administration:** Duong Mylinh, Salim Yusuf, Wei Li, Bo Hu

**Resources:** Salim Yusuf

**Supervision:** Wei Li, Lap Ah Tse

**Validation:** Zhiguang Liu

**Visualization:** Qiujing Cai, Mengxin Chen, Biyan Wang

**Writing - original draft:** Mengya Li, Yang Li, Lap Ah Tse

**Writing – review & editing:** Duong Mylinh, Qiujing Cai, Salim Yusuf, Wei Li, Lap Ah Tse, Zhiguang Liu, Bo Hu

## Acknowledgements

We are thankful for all PURE Project Office Staff, National Coordinators, investigators and the participants of this study.

## PURE-China Project Office Staff, National Coordinators, Investigators

Liu Lisheng*, Li Wei*, Chen Chunming, Zhao Wenhua, Hu Bo, Zhang Hongye, Zhu Jun, Liang Yan, Song Haiqing, Xiong Rong, Li Shujuan, Wang Jiying, Yu Litian, Yang Yanmin, Yang Jun, Bo Jian, Sun Yi, Yin Lu, Deng Qing, Cheng Xiaoru, Wang Yang, Wang Xingyu, He Xinye, Jia Xuan, Zhang Li, Liu Xiaoyun, Han Guoliang, Chen Hui, Liu Xu, Gao Nan, Bai Xiulin, Ren Li, Zhang Jixian, Wang Yue, Tian Bingxue, Ren Meixi, Wang Bing, Tao Xiaoqing, Zhang Yifan, Shao Tianjing, Wang Mengyao, Li Zhe, Dai Xi, Ren Yingjuan, Li Ling, Tao Zhengqing, Gao Liuning, Song Rui, Cao Zhuangni, Yao Chenrui, Deng Guojiao, Liu Xiehui, Wang Yueming, Liu Zhiguang, LA Tse Shelly, Gu Hongqiu, Yan Ruohua, Peng Yaguang, Wang Chuangshi, Li Sidong, Liu Weida, Xia Yanjie, Hao Jun, Zhu Yingxuan, Li Mengya, Lang Xinyue, Li Xiaocong, Yan Minghai, Huang Yilin, Chen Feilong, Li Qi, Danzeng Chilie, Li Qize, Hu Lingqian, Chen Mengxin, Wang Biyan, Liu Xin, Lan Lei, Wang Duoer, Lin Xiaoying, Hou Yan, Zhang Liangqing, Wang Junying, Liu Guoqing, Liao Xiaoyang, Zhao Qian, Xu Guofan, Qing Tao, Zhang Xiaolin, Li Dong, Chen Di, Jin Hui, Tian Jiwen, Zhang Peng, Zhi Yahong, Yuan Shenghua, Fu Xiaoming, Li Ning, Zhang Li, Lei Rensheng, Hu Lihua, Tang Xincheng, Fu Minfan, Wang Qiuyuan, Fu Xiaoli, Liu Yu, Xing Xiaojie, Yang Youzhu, Xu Wenqiang, Hai Yan, Ma Yuanting, Wang Yali, Ma Haibin, Zhao Shenghu, Pan Yingzi, Xiang Quanyong, Tang Jinhua, Liu Zhengrong, Qiang Deren, Qian Zhenzhen, Zhang Yunfei, Xu Zhengting, Han Aiying, Chen Shimei, Yang Jinkui, Liu Chunmei, He Guomin, Liu Huaxing, Wu Xinbin, Ai Hanyue, Resalaiti Aobulikasimu, Wang Hui, Aideer Aili, Ayoufumiti Wula, Lu Fanghong, Su Jing, Zhao Yingxin, Yang Jianmin; Chen Di, Jin Hui, Tian Jiwen, Cheng Xiaoguang, Peng Tao, Li Yin dong, Li Changqing, You Kai, Chen Peng.

## Research Ethics and Patient Consent Ethical approval

The project of “The Prospective Urban and Rural Health Evaluation Study: PURE” is a global study, which was approved on both ethical and scientific grounds by McMaster University Research Ethics Board on July 7, 2003. The study protocol was also approved by the ethics committees of the Fuwai Hospital (NO.2020-1313) of Chinese Academy of Medical Sciences and Beijing Hypertension League Institute. Informed consent was obtained from all participants prior to data collection.

## Abbreviations

AO: airflow obstruction
BMI: body mass index
CI: confidence interval
COPD: chronic obstructive pulmonary disease
CVD: cardiovascular diseases
FEV_1_: forced expiratory volume in one second
FVC: forced vital capacity
HR: hazard ratios
PRISm: preserved ratio impaired spirometry
PURE: Prospective Urban Rural Epidemiology

