## Supplemental Table S1 for "Association between Preserved Ratio Impaired Spirometry with mortality and long-term cardiovascular outcomes in Chinese adults"

**Association between** **Preserved Ratio Impaired Spirometry (PRISm) with Mortality and Long-term Cardiovascular Outcomes in Chinese Adults: Results from the Prospective Urban Rural Epidemiology (PURE) Study**

**Mengya Li1ϯ, Yang Li2ϯ,1 , Duong Mylinh3, Qiujing Cai1, Mengxin Chen1, Deren Qiang4, Biyan Wang1, Sumathy Rangarajan3, Kai You5, Jiying Li6, Salim Yusuf3, Wei Li1, Lap Ah Tse8*,** **Zhiguang Liu7*, Bo Hu1* on behalf of PURE-China Investigators**

**Supplementary Appendix A: Supplementary Methods**

**Appendix A1: PURE Study Participant Selection Methodology**

as Excerpted from Teo et al. Am Heart J. 2009 Jul; 158 (1):1-7

**Selection of Communities**

Within each country, urban and rural communities were selected based on broad guidelines (see Guidelines for Selection of Countries, Communities, Households, and Individuals Recruited to PURE). A common definition for “community” that is applicable globally is difficult to establish. In PURE, a community was defined as a group of people who have common characteristics and reside in a defined geographic area. A city or large town was not usually considered a single community, rather communities from low-, middle-, and high-income areas were selected from sections of the city and the community area defined according to a geographical measure (e.g., a set of contiguous postal code areas or a group of streets or a village). The primary sampling unit for rural areas in many countries was the village. The reason for inclusion of both urban and rural communities is that for many countries, urban and rural environments exhibit distinct characteristics in social and physical environment, and hence, by sampling both, we ensured considerable variation in societal factors across PURE communities.

The number of communities selected in each country varied, with the aim to recruit communities with substantial heterogeneity in social and economic circumstances balanced against the capacity of local investigators to maintain follow-up. In some countries (e.g., India, China, Canada, and Colombia), communities from several states/provinces were included to capture regional diversity, in policy, socioeconomic status, culture, and physical environment.

**Selections of Households and Individuals**

Within each community, sampling was designed to achieve a broadly representative sample of that community of adults aged between 35 and 70 years (see Table below for general recommendations for countries in PURE). The choice of sampling frame within each center was based on both “representativeness” and feasibility of long-term follow-up, following broad study guidelines. Once a community was identified, where possible, common and standardized approaches were applied to the enumeration of households, identification of individuals, recruitment procedures, and data collection.

The method of approaching households differed between regions. For example, in rural areas of India and China, a community announcement was made to the village through contact of a community leader, followed by in-person door-to-door visits of all households. Households were eligible if at least 1 member of the household was between the ages of 35 and 70 years and the household members intended to continue living in their current home for a further 4 years.

For each approach, at least 3 attempts at contact were made. All individuals within these households between 35 and 70 years providing written informed consent were enrolled. When an eligible household or eligible individual in a household refused to participate, demographics and self-reported data about CVD risk factors, education, and history of CVD, cancers and deaths in the households within the 2 previous years were recorded.

To ensure standardization and high data quality, we used a comprehensive operation manual, training workshops, DVDs, regular communication with study personnel and standardized report forms. We entered all data in a customized database programmed with range and consistency checks, which was transmitted, electronically to the Population Health Research Institute in Hamilton (Ontario, Canada) where further quality checks were implemented.

**Guidelines for Selection of Countries, Communities, Households, and Individuals Recruited to PURE**

**Countries**

1. High-income countries, middle-income countries, and low-income countries, with the bulk of the recruitment from low- and middle-income regions.

2. Committed local investigators with experience in recruiting for population studies.

**Communities**

1. Select both urban and rural communities. Use the national definition of the country to determine urban and rural communities.

2. Select rural communities that are isolated (distance of >50 km or lack easy access to commuter transportation) from urban centers. However, consider ability to process bloods samples, e.g., villages in rural developing countries should be within 45-min drive of an appropriate facility.

3. Define community to a geographical area, e.g., using postal codes, catchment area of health service/clinics, census tracts, areas bordered by specific streets or natural borders such as a river bank.

4. Consider feasibility for long-term follow-up, e.g., for urban communities, choose sites that have a stable population such as residential colonies related to specific work sites in developing countries. In rural areas, choose villages that have a stable population. Villages at greater distance from urban centers are less susceptible to large migration to urban centers.

5. Enlist a community organization to facilitate contact with the community, e.g., in urban areas, large employers (government and private), insurance companies, clubs, religious organizations, clinic or hospital service regions. In rural areas, local authorities such as priests or community elders, hospital or clinic, village leader, or local politician.

**Individual**

1. Broadly representative sampling of adults 35 to 70 years within each community unit.

2. Consider feasibility for long-term follow-up when formulating community sampling framework, e.g., small percentage random samples of large communities may be more difficult to follow-up because they are dispersed by distance. In rural areas of developing countries that are not connected by telephone, it may be better to sample entire community (i.e., door-to-door systematic sampling).

3. The method of approach of households/individuals may differ between sites. In MIC and HIC, mail, followed up by phone contact may be the practical first means of contact. In LIC, direct household contact through household visits may be the most appropriate means of first contact.

4. Once recruited, all individuals are invited to a study clinic to complete standardized questionnaires and have a standardized set of measurements.

**Specific recruitment strategies for China are summarized below.**

**Selection of Provinces:** In China, we selected several provinces to capture regional diversity, in policy, socioeconomic status, culture, and physical environment. The choice and number of provinces selected in PURE China reflects a balance between involving a large number of communities in provinces at different economic levels, with substantial heterogeneity in social and economic circumstances and policies, and the feasibility of centres to successfully achieve long-term follow-up. Thus, PURE China study included sites in which investigators are committed to collecting good-quality data over the planned 10-year follow-up period and did not aim for a strict proportionate sampling of the entire country. We included 12 provinces of mainland China to capture regional diversity, policy, socioeconomic status, culture, and physical environment. Based on the national criteria for coordinated regional development according to their socioeconomic development status at the time of inclusion, these provinces were categorized into Eastern (Beijing, Jiangsu, Liaoning, Shandong), Central (Shanxi, Jiangxi), and Western regions (Shaanxi, Sichuan, Yunnan, Inner Mongolia, Qinghai, Xinjiang).

**Selection of Communities:** Within each province, urban and rural communities were selected using pre-specified criteria to recruit participants with an almost 1:1 urban-to-rural proportion. A total of 115 communities were included in the PURE China study.

**Selection of Households and Individuals:** Within each community, households were approached by a community announcement through contact of the community organization, followed by in-person door-to-door visits of all households. Households were eligible if at least 1 member of the household was between the ages of 35 and 70 years and the household members intended to continue living in their current home for a further 4 years. All individuals within these households between 35 and 70 years providing written informed consent were enrolled.

**Appendix A2: Data collection**

Detailed information on data collection in PURE study has been published previously (Teo et al. Am Heart J. 2009 Jul; 158 (1):1-7). Baseline information was collected using standardized questionnaires and physical measurements. The Adult Questionnaire for adult participants collects data on the 9 INTERHEART risk factors (lipids, smoking, hypertension, diabetes, abdominal obesity, psychosocial factors [stress and symptoms of depression], consumption of fruits, and vegetables, consumption of alcohol, and regular physical activity). The physical examination includes 2 measures of resting blood pressure (sitting), anthropometric measures (weight, height, waist, and hip), spirometry (forced expiratory volume in 1 second, forced expiratory vital capacity), electrocardiogram, and hand grip strength (three measurements from both hands). A 10-mL fasting blood sample is collected from all consenting participants. Blood samples are centrifuged and transferred to centralized long-term storage in secure −70°C freezers or large −180°C liquid nitrogen tanks for future biochemical and genetic testing. In addition, detailed information on physical activity and diet is collected using the International Physical Activity Questionnaire (IPAQ) and country-specific semiquantitative Food Frequency Questionnaires (FFQs). Data on PURE study households are collected using the Household Questionnaire obtains information from the household head on house structure, amenities, access to water, and sanitation.

Quality of data collection is maintained through the use of standardized protocols and centralized training. Key staff from each centre attend an initial training session (standardization across regions is provided by Project Office staff attending these training sessions) and in turn, train local staff using centrally created manuals and training videos. These staff are trained, tested on “mock” subjects, and certified. If the variation between the staff compared to the coordinator is unacceptable, the staff member is retrained. All data are entered at each site electronically into a customized database programmed with range and consistency checks and transmitted electronically to the Project Office in Hamilton, Ontario, Canada.

**Appendix A3: Quality control for spirometry test**

Quality of spirometry was maintained through the use of standardized protocols and centralized training. Key staff (regional coordinator) from each center attended an initial training session conducted by the National Center for Cardiovascular Diseases, China every year, and in turn trained local staff using centrally created manuals and training videos. Local staff were tested on mock subjects and certified. If a staff member was unqualified, he/she would be retrained until complying with the standard. All frontline workers should be able to tell whether participants doing spirometry test correctly with maximal effort at the time of measuring. Meanwhile, monthly calibration of spirometer was required, or at any time thought necessarily by the local staff (e.g., before use in extreme temperatures); daily calibration was not demanded, because it was hard to be carried out during fieldwork, especially in some remote areas of China.

Prospective validation of spirometry was done during the follow-up period of the PURE-China Study in November, 2012. A total of 95 participants were randomly selected from an urban community of Beijing, which was one of the largest center in China and was accessible to spirometry laboratories. Concurrent spirometry measurements were obtained using the fieldwork methodology by local staff and following the ATS/ERS standardization [Eur Respir J 2005; 26: 319-338] by respiratory professionals within three hours without any intervening foods, drinks or medications. Results showed that the average differences between lab and field data were 161 mL for FEV1 and 393 mL for FVC. No trend was observed in lab-field difference or its variability with the increase of spirometry values, suggesting the measurements of field spirometry were basically valid.

All data were entered electronically into a customized database with range and consistency checks. The spirometry tracings were inspected by the coordinating center for further quality control. We used 200 mL rather than 150 mL (as recommended by the ATS/ERS standardization) to evaluate the reproducibility of spirometry due to practical concerns. For clinical purposes, ideally reproducibility should be 50-100 mL (i.e., grade A & B) and perhaps 150 mL (grade C). But for epidemiological data, there is no established guideline since very few large population-based surveys has ever been carried out especially in low- and middle-income countries. 200 mL is the upper limit to balance the acceptability of spirometry quality and the cost-effectiveness of data collection.

**Appendix A4:** **Standardized Event Definitions in PURE**

**Prospective Follow-up for Cardiovascular Events and Mortality:** Information on specific events (e.g. death, myocardial infarction, stroke, heart failure, cancer, hospitalizations, new diabetes, injury, tuberculosis, human immunodeficiency viral infections, malaria, pneumonia, asthma, chronic obstructive pulmonary disease) were obtained from participants or their family members (events were reported by the participants if alive or by a relative if the individual had died). Because the PURE study involves urban and rural areas from middle- and low-income countries, supporting documents to confirm the cause of death and/or event varied in degrees of completion and availability. In most of middle- and low-income countries there was no central system of death or event registration. Therefore, information was obtained about prior medical illness and medically certified cause of death where available, and, second, best available information was captured from reliable sources in those instances where medical information was not available in order to be able to arrive at a probable diagnosis or cause of death. Event documentation was based on information from household interviews and medical records, death certificates and other sources. Verbal autopsies were also used to ascertain cause of death in addition to medical records which were reviewed by a health professional. This approach has been used in several studies conducted in middle- and low-income countries.

The following definitions and codes were used to classify CVD events and deaths in the study:

**CVD Definitions and Coding**

FATAL EVENTS

Cardiovascular Death – Definitions

01.00 DEATH DUE TO CARDIOVASCULAR EVENTS

01.10 Sudden unexpected Cardiovascular Death (SCVD)

Without evidence of other cause of death, death that occurred suddenly and unexpectedly (examples: witnessed collapse, persons resuscitated from cardiac arrest who later died) or persons seen alive less than 12 hours prior to discovery of death (example persons found dead in his/her bed).

01.10 SCVD is either definite, probable or possible according to the following characteristics:

| PURE  Adjudication Code | Event Type | Acceptable ICD-10 codes |
| --- | --- | --- |
| 01.11: Definite | One of the following in persons with:   - known cardiovascular disease, or - diabetes with an additional risk factor such as hypertension, smoking, dyslipidemia, micro albuminuria, serum creatinine 50% above upper limit of normal, or - 3 of the above risk factors, or - 2 of the above risk factors in men aged 60 and more and women aged 65 and more | No ICD-10 Code |
| 01.12: Probable | One of the following in persons with:   - diabetes, or - 2 of the above risk factors in men aged less than 60 and in women less than 65, or - one of the above risk factor in men aged 60 and more and in women aged 65 and more, or - typical of chest pain or sudden severe dyspnea of less than 20-minute duration preceding the event |  |
| 01.13: Possible | In persons without risk factor |  |
| *For SCVD, the patient was well or had a stable CVD (example stable angina) when last seen alive. The event of a sudden death occurring during the hospitalization of MI is considered a fatal MI and not sudden death.* | | |

01.3 Fatal Myocardial Infarction (MI)

Symptoms of Myocardial Infarction:

Typical symptoms or suggestive symptoms of MI according to physician are characterized by severe anterior chest pain as tightness, crushing, burning, lasting at least 20 minutes, occurring at rest, or on exertion, that may radiate to the arms or neck or jaw and may be associated with dyspnea, diaphoresis and nausea. However, death associated with nausea and vomiting with or without chest pain not due to another cause may be considered as possible MI if ECG and cardiac markers are not done. These symptoms may have occurred the last month before death.

Fatal myocardial infarction is either definite, probable or possible according to the following characteristics:

| PURE  Adjudication Code | Event Type | Acceptable ICD-10 codes |
| --- | --- | --- |
| 01.31: Definite | 1. Autopsy demonstrating fresh myocardial infarction and/or recent coronary occlusion, or 2. ECG showing new and definite sign of MI (Minnesota code 1-1-1) or 3. Symptoms typical or atypical or inadequately described but attributed to cardiac origin lasting at least 20 minutes and by troponin or cardiac enzymes (CKMB, CK, SGOT, SLDH) above center laboratory ULN 4. ECG with new ischemic changes (new ST elevation/depression or T wave inversion ≥ 2 mm) and by troponin or cardiac enzymes (CKMB, CK, SGOT, SLDH) above center laboratory ULN | I21- I22 |
| 01.32: Probable | 1. ECG with sign of probable MI (Minnesota code 1-2-1), or 2. Typical symptoms lasting at least 20 minutes considered of cardiac origin, with only new ST-T changes (new ST elevation/depression or T wave inversion ≥ 1 but < 2mm) without documented increased cardiac markers or enzyme as in PURE definition 1.31 (above), or 3. Increased cardiac enzymes as in PURE definition 1.31 (above) showing a typical pattern of MI as above without symptoms or significant ECG changes |  |
| 01.33: Possible | 1. ECG with sign of possible MI (Minnesota code 1-3-1) or 2. Typical symptoms or symptoms suggestive of MI according to the physician lasting at least 20 minutes without documented ECG or cardiac marker. |  |

The Minnesota codes for MI is taken from Rose and Blackburn and published in their book “Evaluation Methods of Cardiovascular Disease WHO 1969”.

- Definite  MI is Q/R ratio ≥1/3 and Q duration ≥ 0.03 second in one of the following leads: I, II, V2, 3, 4, 5, 6. (code 1-1-1)
- Probable MI is Q/R ratio ≥1/3 and Q duration between 0.02 and  0.03 second in one of the following leads: I, II, V2, 3, 4, 5, 6. (code 1-2-1)
- Possible  MI is Q/R ratio between 1/5 and 1/3 and Q duration between 0.02 and  0.03 second in one of the following leads: I, II, V2, 3, 4, 5, 6. (code 1-3-1)

01.40 Fatal Stroke

Fatal stroke is either definite or possible according to the following characteristics:

| PURE  Adjudication Code | Event Type | Acceptable ICD-10 codes |
| --- | --- | --- |
| 01.41: Definite | Stroke death is defined as death within 30 days from an acute focal neurological deficit *diagnosed by a physician* and thought to be of vascular origin (without other cause such as brain tumor) with signs and symptoms lasting >= 24 hrs.  Stroke death is also considered if death occurred within 24 hrs. of onset of persisting signs and symptoms, or if there is evidence of a recent stroke on autopsy.  N.B.   - In a subject with a stroke <= 30 days: If death occurred with a pneumonia due to possible aspiration, death will be considered to be due to stroke. - In a subject with a stroke > 30 days: If death occurred with a pneumonia due to possible aspiration, the adjudicator will make a decision according to his/her clinical judgment if death is related to stroke or not. - Subarachnoid hemorrhage death manifested by sudden onset headache with/without focal signs and imaging (CT or MRI) evidence of bleeding primarily in the subarachnoid space is considered a fatal stroke in absence of trauma or brain tumor or malformation - Subdural hematoma death is not considered as a stroke death and may be related to previous trauma or other cause. | I60- I64, I69 |
| 01.43: Possible | Death in a participant with a history of sudden onset of focal neurological deficit of one or more limbs, loss of vision or slurred speech lasting about 24 hours. |  |

01.50 Fatal Congestive Heart Failure

Fatal congestive heart failure is either definite or possible according to the following characteristics:

| PURE  Adjudication Code | Event Type | Acceptable ICD-10 codes |
| --- | --- | --- |
| 01.51: Definite | The diagnosis of congestive heart failure may be an autopsy finding in absence of other cause or requires signs (rales, increased jugular venous pressure or ankle edema) or symptoms (nocturnal paroxysmal dyspnea, dyspnea at rest or ankle edema) of congestive heart failure and one or both of the following:   - radiological signs of pulmonary congestion, - treatment of heart failure with diuretics   *If sudden death occurred in a patient with chronic severe heart failure, it should be adjudicated as fatal congestive heart failure.* | I50 |
| 01.52: Probable | Progressive shortness of breath on lying down or at night, improving on sitting up AND any of the following signs or symptoms: swelling of feet, distension of abdomen, progressive cough in a person with known hypertension or a history of previous MI/angina or other heart disease |  |
| 01.53: Possible | Progressive shortness of breath on lying down or at night, improving on sitting up AND any of the following signs or symptoms: swelling of feet, distension of abdomen, progressive cough |  |

01.60 Death Due to Other Cardiovascular Deaths *(other causes [1.10 to 1.50 above] having been excluded)*

| PURE  Adjudication Code | Event Type | Acceptable ICD-10 codes |
| --- | --- | --- |
| 01.61 | Arterial rupture of aneurysm | I71- I72 |
| 01.62 | Pulmonary embolism  *NOTE: Death associated with pulmonary embolism occurring within 2 weeks after a fracture such as hip, femur should attributed to death due to injury. Refer to Injury, Section 6.0* | I26 |
| 01.63 | Arrhythmic death (A-V block, sustained ventricular tachycardia in absence of other causes) | I44- I45,  I47- I49 |
| 01.64 | Death after invasive cardiovascular intervention: a perioperative death extending to 30 days after coronary or arterial surgical revascularization and to 7 days after a coronary or arterial percutaneous dilatation (angioplasty) with or without a stent or an invasive diagnostic procedure. | I97 |
| 01.65 | Congenital heart disease | Q20-Q28 |
| 01.66 | Heart valve disease (including rheumatic heart disease) | I01, I05- I09,  I34- I37 |
| 01.67 | Endocarditis | I33, I38 |
| 01.68 | Myocarditis | I40 |
| 01.69 | Tamponade (pericarditis) | I30, I31, I32 |
| 01.70 | Other cardiovascular events *(Excluding 1.61 to 1.69 above)*  *Valid ICD-10 codes would include the following:*  *I11, I12, I13, I23, I24, I25, I27, I28, I42, I51, I52, I65-I68, I73, I74, I96, I98, I99 (Refer to ICD-10 Listing for associated definitions for each code)* | Any valid ‘I’ (Cardiovascular) ICD-10 code that can be classified as underlying cause of death, not specified above |

NON-FATAL EVENTS

Cardiovascular Events – Definitions

10.00 NON-FATAL CARDIOVASCULAR EVENTS

10.10 Non-Periprocedural Myocardial Infarction (MI)

MI is considered either definite, probable or possible according to the following characteristics:

| PURE  Adjudication Code | Event Type | Acceptable ICD-10 codes |
| --- | --- | --- |
| 10.11: Definite | 1. ECG showing new and definite sign of MI (Minnesota code 1-1-1) or 2. Symptoms typical or atypical or inadequately described but attributed to cardiac origin lasting at least 20 minutes and by troponin or cardiac enzymes (CKMB, CK, SGOT, SLDH) above center laboratory ULN 3. ECG with new ischemic changes (new ST elevation/depression or T wave inversion ≥ 2 mm) and by troponin or cardiac enzymes (CKMB, CK, SGOT, SLDH) above center laboratory ULN   Please note that increased markers may occur in trauma (CK, AST, myoglobin and CK MB to a lesser degree); renal insufficiency, heart failure, pulmonary embolism (troponin), cardioversion (all) | I21-I22 |
| 10.12: Probable | 1. ECG with new and probable sign of MI (Minnesota code 1-2-1), or 2. Typical symptoms lasting at least 20 minutes considered of cardiac origin, with only new ST-T changes (new ST elevation/depression or T wave inversion ≥ 1 but < 2mm) without documented increased cardiac markers as in PURE definition 10.11 (above), or 3. Increased cardiac enzymes showing a typical pattern of MI as above without symptoms or significant ECG changes. |  |
| 10.13: Possible | 1. ECG with new and possible sign of MI (Minnesota code 1-3-1), or 2. Typical symptoms lasting 20 minutes and more considered to be of cardiac origin without documented ECG or cardiac marker. |  |

10.20 Periprocedural Myocardial Infarction

| PURE  Adjudication Code | Event Type | Acceptable ICD-10 codes |
| --- | --- | --- |
| 10.21: Definite | 1. ECG showing new and definite sign of MI (Minnesota code 1-1-1), or 2. Increased cardiac markers within 48 hours of procedure:  - percutaneous coronary intervention: CKMB should be ≥ 5 X ULN or troponin ≥ 5 X above lower level of necrosis OR > 20% increase in cardiac markers if elevated at the beginning of the procedure in a patient with symptoms suggestive of myocardial ischemia - Coronary surgery: Increased cardiac markers CKMB should be ≥ 10X ULN or troponin ≥ 10X above lower limit of necrosis. | I21-I22 |

The Minnesota codes for MI is taken from Rose and Blackburn and published in their book “Evaluation Methods of Cardiovascular Disease WHO 1969”.

- Definite  MI is Q/R ratio ≥1/3 and Q duration ≥ 0.03 second in one of the following leads: I, II, V2, 3, 4, 5, 6. (code 1-1-1)
- Probable MI is Q/R ratio ≥1/3 and Q duration between 0.02 and  0.03 second in one of the following leads: I, II, V2, 3, 4, 5, 6. (code 1-2-1)
- Possible  MI is Q/R ratio between 1/5 and 1/3 and Q duration between 0.02 and  0.03 second in one of the following leads: I, II, V2, 3, 4, 5, 6. (code 1-3-1)

10.30 Stroke/Transient Ischemic Attack (TIA)

| PURE  Adjudication Code | Event Type | Acceptable ICD-10 codes |
| --- | --- | --- |
| 10.31: Definite | Stroke is defined as an acute focal neurological deficit *diagnosed by a physician* and thought to be of vascular origin (without other case such as brain tumor) with signs and symptoms lasting ≥ 24 hrs.  N.B.   - Subarachnoid hemorrhage manifested by sudden onset headache with/without focal signs and imaging (CT or MRI or lumbar puncture) showing evidence of bleeding primarily in the subarachnoid space is considered a stroke in absence of trauma or brain tumor or malformation - Subdural hematoma is not considered as a stroke and may be related to previous trauma or other cause. | I60-I64, I69 |
| 10.33: Possible | Stroke is possible if there is a history of sudden onset of focal neurological deficit of one or more limbs, loss of vision or slurred speech lasting about 24 hours or more |  |
| 10.34: TIA | The diagnosis of TIA requires the presence of acute focal neurological deficit thought to be of vascular origin with signs and symptoms lasting less than 24 hours | G45 |

10.40 Congestive Heart Failure

| PURE  Adjudication Code | Event Type | Acceptable ICD-10 codes |
| --- | --- | --- |
| 10.41: Definite | The diagnosis of congestive heart failure requires signs (rales, increased jugular venous pressure or ankle edema) or symptoms (nocturnal paroxysmal dyspnea, dyspnea at rest or ankle edema) of congestive heart failure and one or both of the following:   - radiological signs of pulmonary congestion, - Treatment of heart failure with diuretics. | I50 |
| 10.42: Probable | Progressive shortness of breath on lying down or at night, improving on sitting up AND any of the following signs or symptoms: swelling of feet, distension of abdomen, progressive cough in a person with known hypertension or a history of previous MI/angina or other heart disease |  |
| 10.43: Possible | Congestive heart failure is considered possible when there is progressive shortness of breath on lying down or at night, improving on sitting up AND any of the following signs or symptoms: swelling of feet, distension of abdomen, progressive cough |  |

**Coding of Deaths in the PURE Study**

| **Generic Event Codes** | **Description** | **Acceptable FATAL PURE Codes** | | | | **Acceptable ICD 10 Code Ranges** |
| --- | --- | --- | --- | --- | --- | --- |
|  |  | **Definite** | **Probable** | **Possible** | **Codes for which Definite/**  **Probable/**  **Possible not defined** |  |
| **1.00** | **Death Due to Cardiovascular Events** |  |  |  |  |  |
| 1.10 | **Sudden unexpected cardiovascular death** | 1.11 | 1.12 | 1.13 |  | **No ICD Code** |
| 1.30 | **Fatal Myocardial Infarction** | 1.31 | 1.32 | 1.33 |  | **I21-I22** |
| 1.40 | **Fatal Stroke** | 1.41 |  | 1.43 |  | **I60-I64, I69** |
| 1.50 | **Fatal Congestive Heart Failure** | 1.51 | 1.52 | 1.53 |  | **I50** |
| 1.60 | **Other cardiovascular deaths** |  |  |  |  |  |
| - | **-** Arterial rupture of aneurysm |  |  |  | 1.61 | **I71-I72** |
| - | **-** Pulmonary Embolism |  |  |  | 1.62 | **I26** |
| - | **-** Arrhythmic death (A-V block, sustained ventricular tachycardia in absense of other causes) |  |  |  | 1.63 | **I44-I45, I47-I49** |
| - | **-** Death after invasive cardiovascular intervention |  |  |  | 1.64 | **I97** |
| - | **-** Congenital heart disease |  |  |  | 1.65 | **Q20-Q28** |
| - | **-** Heart valve disease (including RHD) |  |  |  | 1.66 | **I01, I05-I09, I34-I37** |
| - | **-** Endocarditis |  |  |  | 1.67 | **I33, I38, I39** |
| - | **-** Myocarditis |  |  |  | 1.68 | **I40, I41** |
| - | **-** Tamponade (Pericarditis) |  |  |  | 1.69 | **I30-I32** |
| - | **-** Other cardiovascular events |  |  |  | 1.70 | **I00, I02, I10-I13, I15, I23-I25, I27, I28, I42, I43, I51, I52, I65-I68, I70, I73, I74, I77-I83, I85-I89, I95, I98, I99** |
| **2.00** | **Death due to most frequent infections** |  |  |  |  |  |
| 2.10 | **Typhoid and Paratyphoid** | 2.11 | 2.12 |  |  | **A01** |
| 2.20 | **Diarrhoea and Gastroenteritis/Dysentery** | 2.21 | 2.22 |  |  | **A00, A02-A09** |
| 2.30 | **Pulmonary Tuberculosis** | 2.31 | 2.32 | 2.33 |  | **A15-A16, A19** |
| - | **Septicaemia** |  |  |  | 2.40 | **A40-A41** |
| 2.50 | **Viral Hepatitis** | 2.51 | 2.52 |  |  | **B15-B19** |
| 2.60 | **AIDS** | 2.61 | 2.62 |  |  | **B20-B24** |
| 2.70 | **Malaria** | 2.71 | 2.72 |  |  | **B50-B54** |
| 2.80 | **Covid-19** | 2.81 | 2.82 |  |  | **U07** |
| - | **Other Infections** |  |  |  | 2.90 | **A17-A18, A20-A39, A42-A99, B00-B09, B25-B49, B55-B99** |
| **3.00** | **Death due to cancer** |  |  |  |  |  |
| - | **Mouth** |  |  |  | 3.01 | **C00-C06, C10, C12, C14** |
| - | **Esophagus** |  |  |  | 3.02 | **C15** |
| - | **Stomach** |  |  |  | 3.03 | **C16** |
| - | **Small Intestine** |  |  |  | 3.04 | **C17** |
| - | **Large intestine including rectum** |  |  |  | 3.05 | **C18-C20** |
| - | **Pancreas** |  |  |  | 3.06 | **C25** |
| - | **Liver** |  |  |  | 3.07 | **C22, C24** |
| - | **Lung/Pleura** |  |  |  | 3.08 | **C33-C34, C38-C39, C45** |
| - | **Breast** |  |  |  | 3.09 | **C50** |
| - | **Prostate** |  |  |  | 3.11 | **C61** |
| - | **Head and Neck** |  |  |  | 3.13 | **C31-C32, C70** |
| - | **Skin** |  |  |  | 3.14 | **C43-C44** |
| - | **Multi-site** |  |  |  | 3.15 | **C97** |
| - | **Other, specify** |  |  |  | 3.16 | **C07-C09, C11, C13, C23, C26, C30, C37, C46-C49, C57-C58, C63, C68-C69, C74-C79, C96, D37-D45, D47, D48** |
| - | **Cervical** |  |  |  | 3.17 | **C53** |
| - | **Uterine/Ovarian** |  |  |  | 3.18 | **C54-C56** |
| - | **Vaginal/vulva** |  |  |  | 3.19 | **C51-C52** |
| - | **Kidney** |  |  |  | 3.20 | **C64, C65** |
| - | **Bladder** |  |  |  | 3.21 | **C66-C67** |
| - | **Anus** |  |  |  | 3.22 | **C21** |
| - | **Testis** |  |  |  | 3.23 | **C62** |
| - | **Penis** |  |  |  | 3.24 | **C60** |
| - | **Brain/spinal cord** |  |  |  | 3.25 | **C71-C72** |
| - | **Leukemia** |  |  |  | 3.26 | **C91-C95, D46** |
| - | **Lymphoma** |  |  |  | 3.27 | **C81-C86, C88** |
| - | **Multiple Myeloma** |  |  |  | 3.28 | **C90** |
| - | **Musculoskeletal (muscle, bones, tendons, ligaments, joints, cartilage)** |  |  |  | 3.29 | **C40-C41** |
| - | **Thyroid** |  |  |  | 3.30 | **C73** |
| - | **Unknown site** |  |  |  | 3.40 | **C80** |
| **4.00** | **4.00 Death due to diseases of the respiratory system** |  |  |  |  |  |
| 4.10 | **Pneumonia** | 4.11 | 4.12 | 4.13 |  | **J12-J18** |
| 4.30 | **Asthma** | 4.31 | 4.32 | 4.33 |  | **J45-J46** |
| 4.40 | **COPD (Obstructive airways disease)** | 4.41 | 4.42 | 4.43 |  | **J44** |
| - | **Other respiratory diseases** |  |  |  | 4.90 | **J00-J11, J20-J43, J47-J99, Q30-Q34** |
| **5.00** | **Death related to pregnancy/delivery/puerperium** |  |  |  |  |  |
| - | **Death related to pregnancy/delivery/ puerperium (Direct obstetrical causes)** |  |  |  | 5.00 | **O00-O99** |
| **6.00** | **Death due to Injury** |  |  |  |  |  |
| - | **Death due to Injury** |  |  |  | 6.00 | **V01-V99, W00-W99, X00-X99, Y00-Y98** |
| **7.00** | **Death due to other causes** |  |  |  |  |  |
| - | **Diseases of Nervous system** |  |  |  | 7.10 | **G00-G99, Q00-Q07** |
| - | **Diseases of Digestive System** |  |  |  | 7.20 | **K00-K93, Q39-Q45** |
| - | **Diseases of genito-urinary system** |  |  |  | 7.30 | **N00-N99, Q50-Q64** |
| - | **Other diseases, specify** |  |  |  | 7.50 | **D50-D89, E00-E90, F00-F99, H58, H59, H94, H95, L00-L99, M00-M99, Q35-Q38, Q65-Q99** |
| **8.00** | **Death due to unspecified/presumed cause** |  |  |  |  |  |
| - | **Presumed cardiovascular death** |  |  |  | 8.10 | **No ICD Code** |
| - | **Presumed cancer death** |  |  |  | 8.20 | **No ICD Code** |
| - | **Unknown death** |  |  |  | 8.30 | **R99** |

**Supplementary Appendix B: Tables and Figures**

**Supplementary eTable1: Definitions, measurements, and categorizations of covariates**

| **Variables** | **Definition or method of measurement** |
| --- | --- |
| **Demographic factors:** |  |
| Age | Age at baseline. |
| Sex | Male and female. |
| **Socioeconomic status:** |  |
| Location | Use the national definition of the country to determine urban and rural communities. rural communities that are isolated (distance of >50 km or lack easy access to commuter transportation) from urban centres. However, consider ability to process bloods samples, e.g., villages in rural developing countries should be within 45-min drive of an appropriate facility |
| Education | Education was self-reported, and classified as low (primary education level or less), intermediate (secondary school education) or high (college, trade school, or university education). |
| Occupation | Occupation was self-reported, and classified as professionals/managers, skilled workers, unskilled workers, and homemakers. |
| Household wealth index | Household wealth index was developed based on household possessions by 14 assets and land from the PURE baseline household questionnaire. The items included: electricity, car, computer, television, motorbike, livestock, fridge, other 4 wheelers, washing machine, stereo, bicycle, kitchen processor, telephone, land, and kitchen window from the PURE questionnaire and classified into three categories using the tertile value. |
| **Lifestyle behaviors:** |  |
| Tobacco use | Self-reported tobacco consumption using a standard tobacco use frequency questionnaire, categorized as never, former or current. |
| Pack-years | Self-reported pack-years were defined as years smoked multiplied by the daily number of smoked cigarettes divided by 20. |
| Second-hand smoking | Self-reported second-hand smoking exposure or not. |
| Alcohol consumption | Self-reported alcohol consumption using a standard alcohol consumption frequency questionnaire. Consumption was categorized as former, never and current. Current consumption is further categorized as low (<=7 drinks/week), moderate (8-14 drinks/week in women or 8-21 drinks/week in men), or high consumption (> 14 drinks/week in women or >21 drinks/week in men). |
| Physical activity | Physical activity was measured using the International Physical Activity Questionnaire, and classified as low (<600 metabolic equivalents [MET] × minutes per week or <150 minutes per week of moderate intensity physical activity), moderate (600–3000 MET × minutes or 150–750 minutes per week) or high (>3000 MET × minutes or >750 minutes per week). |
| **Physical examinations:** |  |
| Body mass index | Body mass was measured to the nearest 0·1 kg of body mass by using a precision health scale manufactured by A and D Company of Tokyo, Japan. Stature (height) measurements were recorded to the nearest 0·1 cm using a calibrated stadiometer. Body mass index was calculated from body mass in kilogram divided by height in metre squared (kg/m^2^). Body mass index was categorised according to the World Health Organization (WHO) cut points. Body mass index < 18·5 kg/m² = underweight; 18·5 kg/m² – 24·99 kg/ m² = normal; BMI 25 kg/m² – 29·99 kg/m² = overweight and BMI ≥ 30 kg/m² = obese. |
| Waist-hip ratio | Waist and hip circumference measurements were recorded to the nearest 0.1 cm by using a non-stretchable standard Lufkin tape measure manufactured by Cooper Tools of Apex, North Carolina, United States. |
| Handgrip strength | Handgrip strength was measured using a hand-held model (T.K.K.54010 Takei) dynamometer with the participants in a seated position with the elbow of the dominant hand to be tested flexed at a 90º degree angle. The dominant hand was tested twice and both values were recorded in kilogram. Following the procedures prescribed by the World Health Organization, the given steps were followed to take the HGS measurements. |
| **Metabolic factors:** |  |
| Hypertension | Hypertension was defined by self-report of the disease, taking BP-lowering treatment, or an average SBP at least 140 mmHg and/or an average DBP at least 90 mmHg measured during the visit. |
| Diabetes | Diabetes was defined as either a fasting glucose > 7 mmol/dl or self-reported history of diabetes, on treatment for diabetes. |
| Total cholesterol | Total cholesterol was measured using fasting lipid values. |
| Triglycerides | Triglycerides was measured using fasting lipid values. |
| **Medical history:** |  |
| COPD | Self-reported with history of COPD. |
| Asthma | Self-reported with history of Asthma. |
| CVDs | Self-reported with history of CVDs (including coronary artery diseases, stroke, heart failure and other heart diseases). |
| **Air pollution exposures:** |  |
| Household air pollution from solid fuel use for cooking | Self-reported primary use of solid fuels (i.e. charcoal, coal, wood, agriculture/crop, animal dung, shrub/grass), kerosene, gas or electricity for cooking. |
| Ambient particulate matter 2.5 concentration | Annual PM2.5 concentrations were estimated based on satellite and fixed monitoring data by using a geographically weighted regression model with a resolution of 1 km * 1 km. To estimate aerosol optical depth, multiple satellite products were analyzed and combined with data from the Sun Photometer and GEOS-Chem simulations. These estimates were used to predict ground-level annual PM2.5 concentrations for each community of PURE study. All people living in the same community were assigned the same concentration of PM2.5. |

Abbreviations: BMI: body mass index; BP: blood pressure; COPD: chronic obstructive pulmonary disease; CVD: cardiovascular disease; DBP: diastolic blood pressure; MET: metabolic equivalent; PM 2.5: particulate matter 2.5; SBP: systolic blood pressure; WHO: World Health Organization

**Supplementary eTable2: Hazard ratios (95% CI) comparing PRISm to AO**

| **Outcomes** | **Model 1** | **Model 2** |
| --- | --- | --- |
| All-cause mortality | 1.10 (0.95, 1.26) | 1.10 (0.94, 1.28) |
| Cardiovascular mortality | 1.03 (0.81, 1.30) | 0.89 (0.69, 1.14) |
| Major CVDs | 1.22 (1.08, 1.38) | 1.06 (0.93, 1.21) |
| Myocardial infarction | 1.16 (0.92, 1.47) | 1.00 (0.77, 1.28) |
| Stroke | 1.22 (1.06, 1.42) | 1.06 (0.91, 1.24) |
| Heart failure | 1.42 (0.94, 2.14) | 1.38 (0.88, 2.16) |

Model 1: adjust for age, sex, location, and random effect of centres·

Model 2: adjust for age, sex, location, BMI, waist to hip ratio, education, occupation, household wealth index, tobacco use, pack-years of smoking, second-hand smoking exposure, alcohol consumption, physical activity, handgrip strength, household cooking fuel, PM 2.5, total cholesterol, triglycerides, medical histories of COPD, asthma, cardiovascular diseases, hypertension, diabetes, regular medication intake, and random effect of centers·

**Supplementary eTable3: Hazard ratios (95% CI) from sensitivity analyses for outcomes with lung function category**

|  | **Normal spirometry** | **PRISm** | **Airflow obstruction** |
| --- | --- | --- | --- |
| **All-cause mortality** |  |  |  |
| Using LLN thresholds for FEV1 and FEV1/FVC | 1·00 (ref) | 1·42 (1·29, 1·57) | 1·30 (1·12, 1·50) |
| Excluding participants with COPD and asthma at baseline ^1^ | 1·00 (ref) | 1·43 (1·29, 1·58) | 1·30 (1·12, 1·50) |
| Excluding participants with CVDs at baseline ^2^ | 1·00 (ref) | 1·43 (1·28, 1·59) | 1·29 (1·10, 1·51) |
| Excluding participants who had events in the first two years of follow-up ^3^ | 1·00 (ref) | 1·42 (1·29, 1·57) | 1·30 (1·12, 1·50) |
| Excluding non-smoker from all three groups, and excluding participants with FVC<0·8 pred in normal spirometry group ^4^ | 1·00 (ref) | 1·44 (1·21, 1·71) | 1·38 (1·09, 1·73) |
| Excluding participants with grip strength in the lowest quartile ^5^ | 1·00 (ref) | 1·38 (1·23, 1·56) | 1·37 (1·16, 1·63) |
| Further adjusting for FEV1 | 1·00 (ref) | 1·15 (1·00, 1·31) | 1·01 (0·84, 1·21) |
| Further adjusting for respiratory exacerbations | 1·00 (ref) | 1.42 (1.28, 1.57) | 1.28 (1.10, 1.48) |
| **Major CVDs** |  |  |  |
| Using LLN thresholds for FEV1 and FEV1/FVC | 1·00 (ref) | 1·34 (1·13, 1·59) | 1·51 (1·19, 1·92) |
| Excluding participants with COPD and asthma at baseline ^1^ | 1·00 (ref) | 1·36 (1·15, 1·61) | 1·54 (1·21, 1·96) |
| Excluding participants with CVDs at baseline ^2^ | 1·00 (ref) | 1·25 (1·02, 1·52) | 1·58 (1·21, 2·07) |
| Excluding participants who had events in the first two years of follow-up ^3^ | 1·00 (ref) | 1·34 (1·13, 1·59) | 1·51 (1·19, 1·92) |
| Excluding non-smoker from all three groups, and excluding participants with FVC<0·8 pred in normal spirometry group ^4^ | 1·00 (ref) | 1·29 (1·11, 1·51) | 1·21 (0·97, 1·51) |
| Excluding participants with grip strength in the lowest quartile ^5^ | 1·00 (ref) | 1·13 (1·03, 1·24) | 1·08 (0·94, 1·25) |
| Further adjusting for FEV1 | 1·00 (ref) | 1·12 (1·00, 1·24) | 1·04 (0·90, 1·20) |
| Further adjusting for respiratory exacerbations | 1·00 (ref) | 1.36 (1.14, 1.61) | 1.55 (1.22, 1.98) |
| **Cardiovascular mortality** |  |  |  |
| Using LLN thresholds for FEV1 and FEV1/FVC | 1·00 (ref) | 1·16 (1·07, 1·25) | 1·08 (0·96, 1·23) |
| Excluding participants with COPD and asthma at baseline ^1^ | 1·00 (ref) | 1·16 (1·07, 1·25) | 1·09 (0·96, 1·23) |
| Excluding participants with CVDs at baseline ^2^ | 1·00 (ref) | 1·10 (1·01, 1·21) | 1·08 (0·94, 1·23) |
| Excluding participants who had events in the first two years of follow-up ^3^ | 1·00 (ref) | 1·16 (1·07, 1·25) | 1·08 (0·96, 1·23) |
| Excluding non-smoker from all three groups, and excluding participants with FVC<0·8 pred in normal spirometry group ^4^ | 1·00 (ref) | 1·19 (0·88, 1·62) | 1·62 (1·09, 2·41) |
| Excluding participants with grip strength in the lowest quartile ^5^ | 1·00 (ref) | 1·27 (1·03, 1·57) | 1·49 (1·11, 2·00) |
| Further adjusting for FEV1 | 1·00 (ref) | 1·03 (0·82, 1·29) | 1·11 (0·82, 1·50) |
| Further adjusting for respiratory exacerbations | 1·00 (ref) | 1.15 (1.06, 1.25) | 1.07 (0.95, 1.21) |
| **Myocardial infarction** |  |  |  |
| Using LLN thresholds for FEV1 and FEV1/FVC | 1·00 (ref) | 1·34 (1·16, 1·56) | 1·35 (1·06, 1·72) |
| Excluding participants with COPD and asthma at baseline ^1^ | 1·00 (ref) | 1·34 (1·15, 1·56) | 1·33 (1·04, 1·69) |
| Excluding participants with CVDs at baseline ^2^ | 1·00 (ref) | 1·30 (1·10, 1·55) | 1·36 (1·03, 1·79) |
| Excluding participants who had events in the first two years of follow-up ^3^ | 1·00 (ref) | 1·34 (1·16, 1·56) | 1·35 (1·06, 1·72) |
| Excluding non-smoker from all three groups, and excluding participants with FVC<0·8 pred in normal spirometry group ^4^ | 1·00 (ref) | 1·33 (1·00, 1·76) | 1·36 (0·89, 2·09) |
| Excluding participants with grip strength in the lowest quartile ^5^ | 1·00 (ref) | 1·26 (1·05, 1·51) | 1·10 (0·80, 1·50) |
| Further adjusting for FEV1 | 1·00 (ref) | 1·33 (1·09, 1·62) | 1·33 (1·00, 1·77) |
| Further adjusting for respiratory exacerbations | 1·00 (ref) | 1.34 (1.15, 1.56) | 1.33 (1.05, 1.70) |
| **Stroke** |  |  |  |
| Using LLN thresholds for FEV1 and FEV1/FVC | 1·00 (ref) | 1·04 (0·94, 1·15) | 0·98 (0·84, 1·13) |
| Excluding participants with COPD and asthma at baseline ^1^ | 1·00 (ref) | 1·04 (0·94, 1·15) | 0·98 (0·85, 1·13) |
| Excluding participants with CVDs at baseline ^2^ | 1·00 (ref) | 1·01 (0·91, 1·12) | 1·00 (0·85, 1·17) |
| Excluding participants who had events in the first two years of follow-up ^3^ | 1·00 (ref) | 1·04 (0·94, 1·15) | 0·98 (0·84, 1·13) |
| Excluding non-smoker from all three groups, and excluding participants with FVC<0·8 pred in normal spirometry group ^4^ | 1·00 (ref) | 1·20 (1·00, 1·44) | 1·04 (0·79, 1·35) |
| Excluding participants with grip strength in the lowest quartile ^5^ | 1·00 (ref) | 1·04 (0·93, 1·16) | 1·03 (0·87, 1·22) |
| Further adjusting for FEV1 | 1·00 (ref) | 1·01 (0·89, 1·15) | 0·95 (0·80, 1·13) |
| Further adjusting for respiratory exacerbations | 1·00 (ref) | 1.04 (0.94, 1.14) | 0.97 (0.84, 1.12) |
| **Heart failure** |  |  |  |
| Using LLN thresholds for FEV1 and FEV1/FVC | 1·00 (ref) | 2·01 (1·46, 2·78) | 1·45 (0·93, 2·25) |
| Excluding participants with COPD and asthma at baseline ^1^ | 1·00 (ref) | 2·04 (1·47, 2·83) | 1·53 (0·98, 2·38) |
| Excluding participants with CVDs at baseline ^2^ | 1·00 (ref) | 1·76 (1·20, 2·58) | 1·27 (0·75, 2·15) |
| Excluding participants who had events in the first two years of follow-up ^3^ | 1·00 (ref) | 2·01 (1·46, 2·78) | 1·45 (0·93, 2·25) |
| Excluding non-smoker from all three groups, and excluding participants with FVC<0·8 pred in normal spirometry group ^4^ | 1·00 (ref) | 2·48 (1·29, 4·78) | 2·38 (1·00, 5·65) |
| Excluding participants with grip strength in the lowest quartile ^5^ | 1·00 (ref) | 1·94 (1·30, 2·90) | 1·44 (0·82, 2·52) |
| Further adjusting for baseline FEV1 | 1·00 (ref) | 1·61 (1·06, 2·47) | 1·12 (0·65, 1·94) |
| Further adjusting for respiratory exacerbations | 1·00 (ref) | 1.98 (1.43, 2.74) | 1.41 (0.91, 2.20) |

Abbreviations: BMI: body mass index; COPD: chronic obstructive pulmonary disease; CVD: cardiovascular disease; FEV1, forced expiratory volume in 1 s; FVC, forced vital capacity; LLN, lower limit normal; PM 2.5: particulate matter 2.5; PRISm, Preserved Ratio Impaired Spirometry.

Full model adjusted for age, sex, location, BMI, waist to hip ratio, education, occupation, household wealth index, tobacco use, pack-years of smoking, second-hand smoking exposure, alcohol consumption, physical activity, handgrip strength, household cooking fuel, PM 2.5, total cholesterol, triglycerides, medical histories of COPD, asthma, cardiovascular diseases, hypertension, diabetes, regular medication intake, and random effect of centers in this analysis.

1 Excluding 189 participants with COPD and asthma at baseline;

2 Excluding 3,112 participants with CVDs at baseline;

3 Excluding 427 participants who had events in the first two years of follow-up;

4 Excluding 10894 participants who are current or former smokers, and 135 participants with FVC<0.8 pred in normal spirometry group;

5 Excluding 9632 participants with grip strength in the lowest quartile.

**Supplementary Legends**

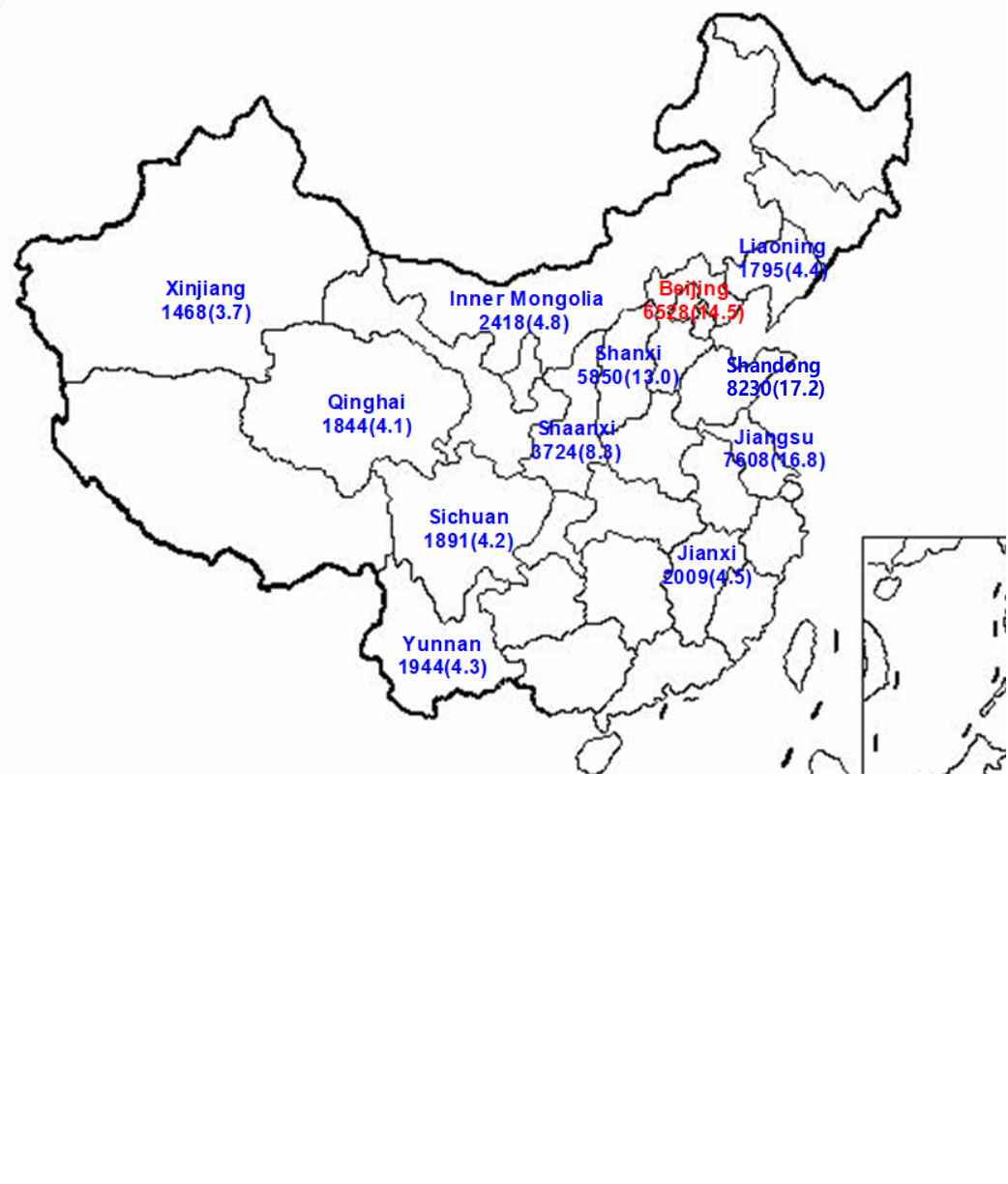

**Supplementary eFigure1: Communities in China participating in the PURE study**

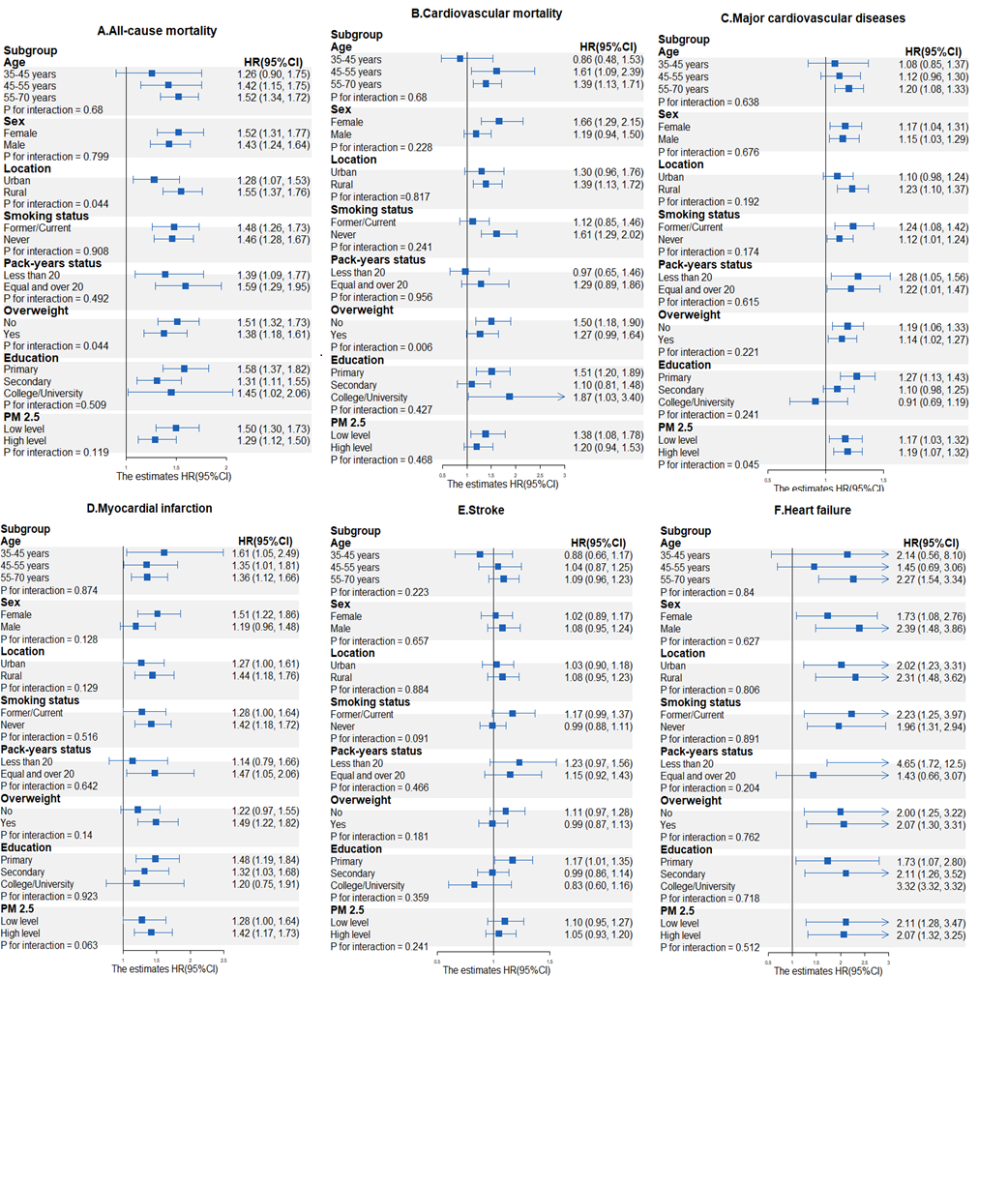

**Supplementary eFigure2: Forest plots of subgroup analysis comparing PRISm with normal group**

Abbreviations: PM 2.5: particulate matter 2.5; PRISm, Preserved Ratio Impaired Spirometry.

Full model adjusted for age, sex, location, BMI, waist to hip ratio, education, occupation, household wealth index, tobacco use, pack-years of smoking, second-hand smoking exposure, alcohol consumption, physical activity, handgrip strength, household cooking fuel, PM 2.5, total cholesterol, triglycerides, medical histories of COPD, asthma, cardiovascular diseases, hypertension, diabetes, regular medication intake, and random effect of centers in this analysis.
